# Country differences in hospitalisation, length of stay, admission to Intensive Care Units, and mortality due to SARS-CoV-2 infection at the end of the first wave in Europe: a rapid review of available literature

**DOI:** 10.1101/2020.05.12.20099473

**Authors:** Elizabeth A Lane, Damien J Barrett, Miriam Casey, Conor G. McAloon, Áine B. Collins, Kevin Hunt, Andrew W. Byrne, David McEvoy, Ann Barber, John Griffin, Patrick Wall, Simon J. More

**Author notes:** Corresponding author: Elizabeth A Lane, Tel: Int-353-01-5058706; Mobile: Int-353-87-2433849.

## Abstract

**Objectives:** Coronavirus disease (COVID-19) caused by the SARS-CoV-2 virus is spreading rapidly worldwide and threatening the collapse of national health care systems. The development of effective resource models are critical for long term health planning. The aim was to evaluate the available literature, to consider parameters affecting hospital resources, to effectively guide health policy and planning for future waves of infection.

**Design:** A detailed search of the literature, using Google Scholar, PubMED, MedRxiv and BioRxiv, was conducted for the time period 1^st^ Dec 2019 to 31^st^ May 2020; using appropriate keywords: resultant articles were scrutinised in detail, and appraised for reported data pertaining to hospitalization and hospital length of stay (LOS).

**Results:** Disease presentation was described in China; 81 % mild, 14 % moderate and 5 % severe. The experience, thus far, in Europe and the USA are suggestive of a higher degree of severity. Initial reports suggest high hospitalisation and ICU admittance rates. More recent reports from the European Centre for Disease Prevention and Control (ECDC) lower this estimation. Perhaps the relative age, the level of pre-existing conditions, and other health factors may be contributors to differences. Data from Irish cases suggest hospitalisation rate may be lower in parts of Europe and time dependent. Hospital LOS is described in 55 articles, with median lengths of stay between 3 and 52 days. The evidence regarding the LOS in ICU is reported in 31 studies, 26 deemed relevant. The majority of studies report ICU LOS between 7 to 11 days. Many of these studies are likely skewed towards shorter stay due to study cut-off dates. Indications based on ICU LOS reported for patients continuing care suggest median ICU stay will progressively increase.

**Conclusions:** These parameter estimates are key to the development of an effective health care resource model. Based on our appraisal of the literature, is it essential that Europe manages mitigation measures to ensure that hospital and ICU capacity does not become overwhelmed to manage COVID-19 in subsequent infection waves.

**Strengths and limitations of this study:**

- The study provides timely information on the differences in hospitalisation, length of stay and ICU length of stay due to COVID-19 in a number of countries worldwide at the end of wave one in Europe;
- This rapid review builds on a previously available review paper that reported length of stay in the early phase of the pandemic; many more studies outlining length of stay, and in particular, ICU length of stay, are now available;
- This rapid review reports on study mortality rate giving an interesting insight into differences across countries and continents;
- Limitations associated with any rapid review are pertinent to this study; a narrow aim was set, and the sources of the literature may be limited by the time-limited constraint of gathering relevant literature; and a number of articles available were in pre-print form and only undergoing peer review; and
- This rapid review provides evidence-based estimates of Hospital and ICU length of stay due to COVID-19 infection across a number of countries to steer policy and provide parameter estimates for utilisation within a hospital resource model as preparations are made for subsequent waves of infection.

## INTRODUCTION

Coronavirus disease (COVID-19) caused by the SARS-CoV-2 virus is spreading rapidly across the world, threatening the collapse of many national health care systems and exerting a grave impact on the global economy. As of 31^st^ May 2020, data compiled from the European Centre for Disease Prevention and Control (ECDC) [1] reported 6,028,135 confirmed infections of COVID-19 cases worldwide with 368,944 deaths recorded. The SARS-CoV-2 virus, first reported in December 2019 in Wuhan China, has spread to 213 countries and territories. By 31^st^ May 2020, European countries, including the European Union (EU-27), the UK, Norway, Switzerland and Iceland have reported 1,429,104 cases and 166,421 deaths following infection with SARS-CoV-2 infection [1]

Clinical presentations of COVID-19 range from asymptomatic to life threatening and fatal. In China, 81 % of COVID-19 cases were considered mild, 14 % severe and 5 % critical [2,3]. The experience in European countries has differed, and varied among countries. The eighth update report from the ECDC [4] indicates that up to 30 % of diagnosed COVID-19 cases were hospitalised in some countries. Individual studies from Italy [5,6] indicated a higher proportion were accessing intensive care unit (ICU) care, with 16 and 18 % of all COVID-19 patients being cared for in an ICU setting. However, the European wide analysis report [4] suggests that 2.4 % of cases presented with severe illness requiring respiratory support or ICU care; the most recent report; the ninth update [7] lowers this estimate (median 2 %).

Worldwide, age and co-morbidities have been demonstrated as major risk factors for the development of severe disease [3,4,7-14]. Children are implicated as possible asymptomatic carriers of SARS-CoV-2 virus and a number of articles report both symptomatic and asymptomatic infection in children [3,15-20]. Many, but not all [17], studies indicate a higher proportion of cases are male [4,7-9, 21-24]; and recent risk assessments conducted by the ECDC [4,7] indicates that males are at higher risk of death compared with their female contemporaries. Close contact with a case is a risk factor [2], and the basis of public health advice includes social distancing and staying at home apart from essential activity [4]. Other factors, including smoking [25] have been highlighted as risk factors for severe infection. There are indications too that infection rate may be altered by blood-group [21,26].

The prevention of infection among health care workers is of critical importance to the management of this pandemic. Of serious concern to policymakers is the clear evidence indicating that both healthcare workers and hospitalised persons are at particularly high risk of exposure [2, 27]. Indeed some of the work conducted early in the epidemic reported that hospital-associated transmission of virus was suspected in 41% of patients [28]. Of concern too, many healthcare authorities are bringing retired medics and nurses back into service. The placement of these people considered vulnerable due to their age must be considered. At a national level, the latest work by Stier et al. [29] emphasizes population density, with attack rates increasing with city size.

Preparedness is key and resource matching to clinical requirement essential. Previous work [30] suggests a large variation in availability of critical care bed in the EU / EEA area, ranging from 29.2 in Germany to 4.2 in Portugal for critical care beds per 100,000 population. Perhaps this is key to understanding the difference in coping between different EU countries. Clearly too, differences will exist between cultures, few western governments will seek to use hospitalisation as a tool for quarantine. An comprehensive review on length of hospital stay in the early part of the pandemic was conducted by Rees et al. [31]. Large variations across studies within China (4 to 53 days) and outside China (4 to 21 days) were reported. However the early studies offered limited evidence for length of ICU stay. Rees et al. [31] reports ICU stay in 8 studies, four within and four outside China, and limited by study subjects so early in the pandemic. Many of the early studies reported hospital length of stay (LOS) for groups with a defined outcome, died or discharged, many too indicated a short follow up time. It is key to consider the influence of those continuing in care. As the infection is spreading across the world, this rapid review paper considers parameters affecting hospital resources resulting from infection with SARS-CoV-2 beyond the evidence published from China. The objective is to evaluate the current scientific literature from across the developed world with the overall aim to influence health policy and support evidence-based decision-making as countries in Europe prepare for a second wave of infection. The proportion of patients hospitalized and their outcomes in terms of mortality rate due to infection with SARS-CoV-2 infection is discussed. The proportion of patients who required critical or ICU care is outlined. Time spent hospitalised, both in a general and an ICU setting, is considered. For those who have been admitted to an ICU, the length of time pre-ICU, the length of time in ICU was collated from available literature. These estimates are central to national resource modelling, which are being developed to aid policymakers to ensure optimal resourcing during subsequent infection waves in this pandemic.

## MATERIALS AND METHODS

### Initial study appraisal

This paper follows the Preferred Reporting Items for Systematic Reviews and Meta-Analyses—Extension for Scoping Reviews (PRISMA-ScR) checklist [32]. A comprehensive search of the scientific literature and national and international government reports was conducted by contributing researchers for the time period 1^st^ December 2019 to 20^th^ May 2020. The search engines Google Scholar, PubMED, MedRxiv and BioRxiv were used, using the following keywords: (“Novel coronavirus” OR “SARS-CoV-2” OR “2019-nCoV” OR “COVID-19”) AND (“length of stay” OR “duration of stay” OR “hospital stay” OR “ICU”). No restriction on language was imposed as long as the abstract was available in English. References within these publications were also searched as additional possibilities for inclusion. Inclusion criteria for ‘the proportion of cases hospitalised’ were papers that reported the proportion of hospitalised and proportion of those hospitalised accessing higher level care including critical care and ICU care. Inclusion criteria for LOS included articles that reported length of hospital stay, length of stay in ICU, and length of stay prior to admission to ICU. Each article was assessed briefly for suitability for inclusion in this study. Papers that did not report hospitalisation, length or duration of time in hospital or in ICU were discarded. The latest governmental reports from the Health Protection Surveillance Centre (HPSC), Ireland [33], and from the Intensive Care National Audit and Research Centre (ICNARC), UK [24] were consulted after the 30^th^ May cut-off.

### Study appraisal

All articles and reports that described the proportion of COVID-19 positive cases hospitalised, the proportion of cases that were admitted to higher level care including critical or ICU care, the length of hospital stay or the duration of hospital stay or length of time in critical care or ICU were scrutinised in detail, and appraised for reported data and quality of the scientific evidence. Parameter estimates for the length of stay in non-ICU or in an ICU setting from the relevant articles were recorded and evaluated.

For quality control, studies were (1) selected from search terms outlined above and initially screened by three members of the team (*KH, SM and LL*), with parameters identified and recorded; (*2*) reviewed and supplemented by manual search by *LL*, again with parameters identified and recorded, and (3) the review was then internally reviewed by an additional three members of the team (*SM, DB, MC*), and cross-referenced with other parameter synthesis documents being developed by the group (*all authors*). Details of the resultant list (n = 263), which included published and pre-printed articles, were entered in an Excel file. Some 55 studies relating to hospital LOS and 31 studies or reports relating to LOS in ICU were selected for detailed scrutiny as they each reported summary estimates with a corresponding variance.

### Source data and management

Data tabulating the number of notified cases and deaths for all countries and Territories worldwide for each calendar date was downloaded in Excel format from the European Centre for Disease Prevention and Control [1]. Data was imported into STATA version 14 (STATA Corporation, USA). Sum of cases and deaths were tabulated for European countries of the EU, the UK, Switzerland, Iceland and Norway (n = 31). Cumulative cases and deaths were generated for each calendar day for countries of interest and expressed as a percentage of cumulative deaths to cumulative cases. The crude mortality rate was expressed as a percentage over time, from day of outbreak, defined as the day on which the tenth case was reported, in selected European Countries.

Data regarding the epidemiological situation in Ireland was extracted from the Health Protection Surveillance Centre daily epidemiological reports [33]. Data were entered into Excel, then imported into STATA 14 to generate the trends in cumulative cases, percentage of known cases hospitalised and the percentage of confirmed cases admittance into ICU over time.

## RESULTS

### TIMEPOINT OF THE PANDEMIC

At study cut-off (31MAY2020), the European first wave of COVID-19 infection was ending and government managed lockdown measures were being eased across the EU, the UK, Switzerland, Norway and Iceland. Figure 1. Illustrates the European Centre for Disease Control and Prevention (ECDC) notified COVID-19 cases and deaths in 31 European countries (EU-27, UK, Switzerland, Norway and Iceland), and the seven day moving average during the first wave of the pandemic; while a mean of 33k cases were notified daily in the 31 European countries during the peak of the first wave of COVID-19 infections between 27MAR2020 to 05APR2020; the ten day period beginning 05JUN2020 recorded just mean of 5.5k daily.

**Figure 1.**
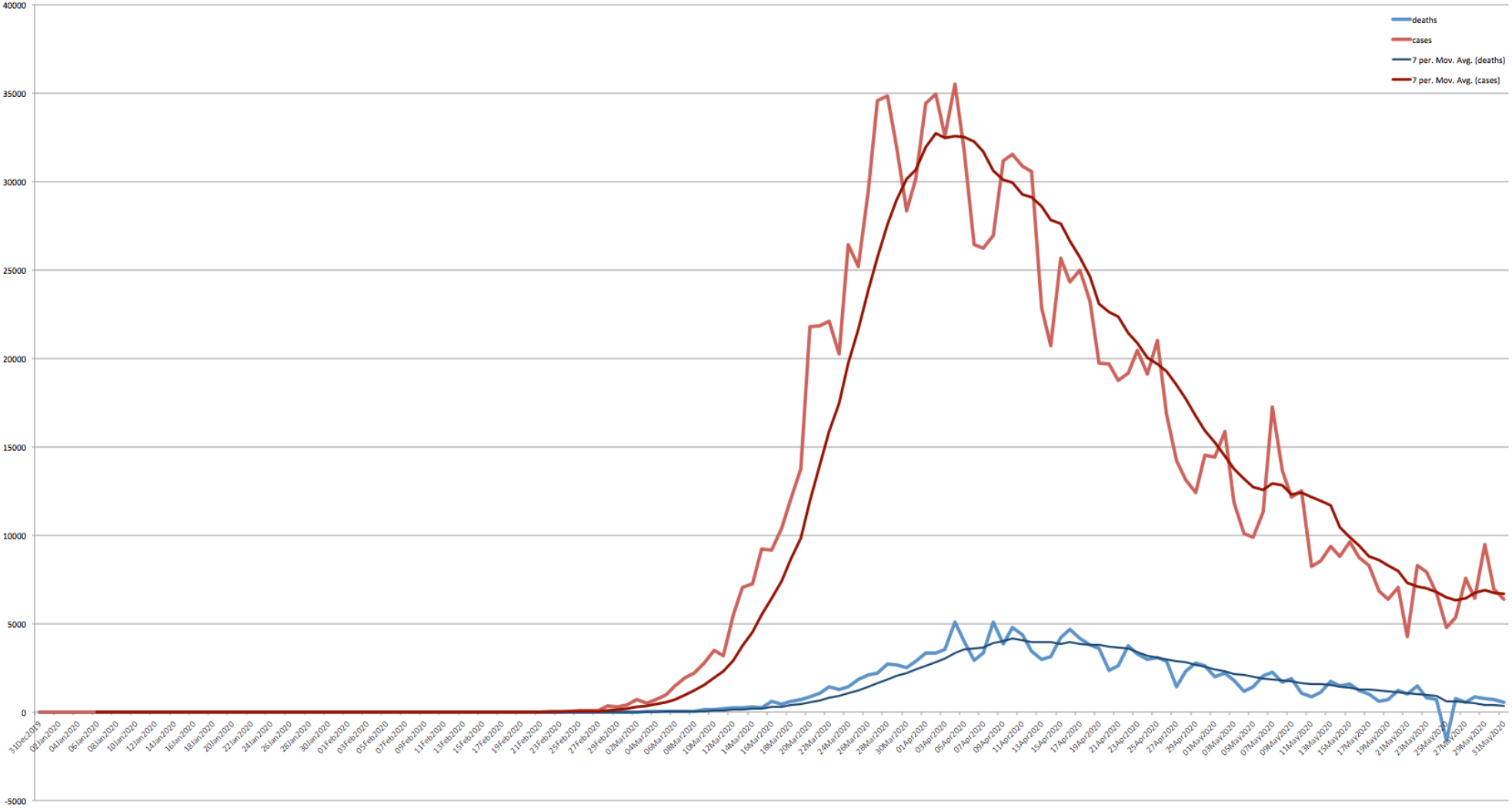
The European Centre for Disease Prevention and Control (ECDC) notified COVID-19 cases and deaths over time in the EU-27, UK, Switzerland, Norway and Iceland. Data tabulating the notified cases and deaths from all countries is available from the European Centre for Disease Control and Prevention; https://www.ecdc.europa.eu/en/publications-data/download-todays-data-geographic-distribution-covid-19-cases-worldwide

### HOSPITALISATION RATE

A summary of available scientific literature from the outbreaks worldwide relating to the proportion of COVID-19 cases requiring hospitalisation and the need for stepped up medical care, including critical care and ICU, by country, and hospital length of stay (LOS) where reported is presented in Table 1. Data from 81 studies or reports are presented, of which 55 describe to hospital LOS.

**Table 1.**
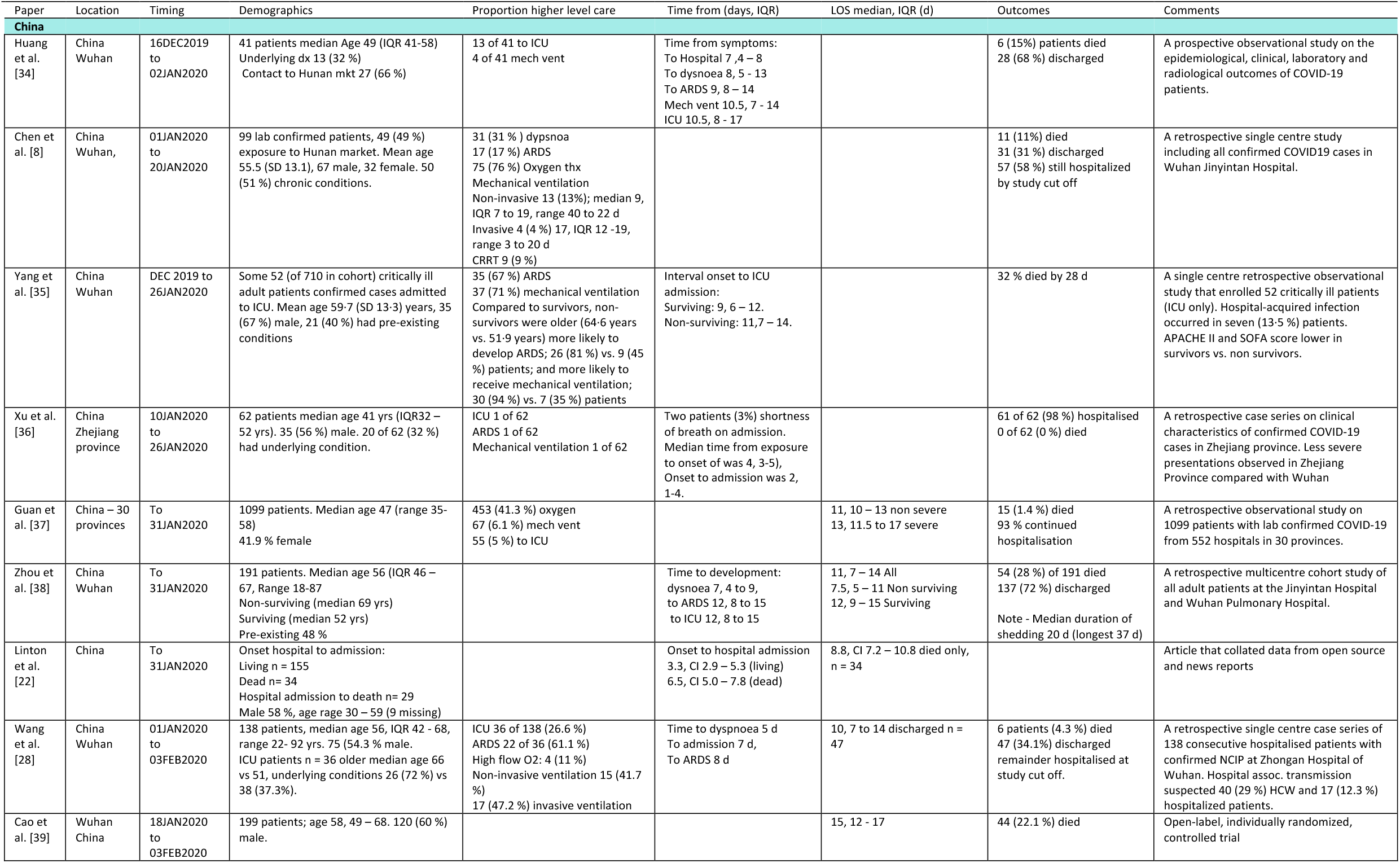
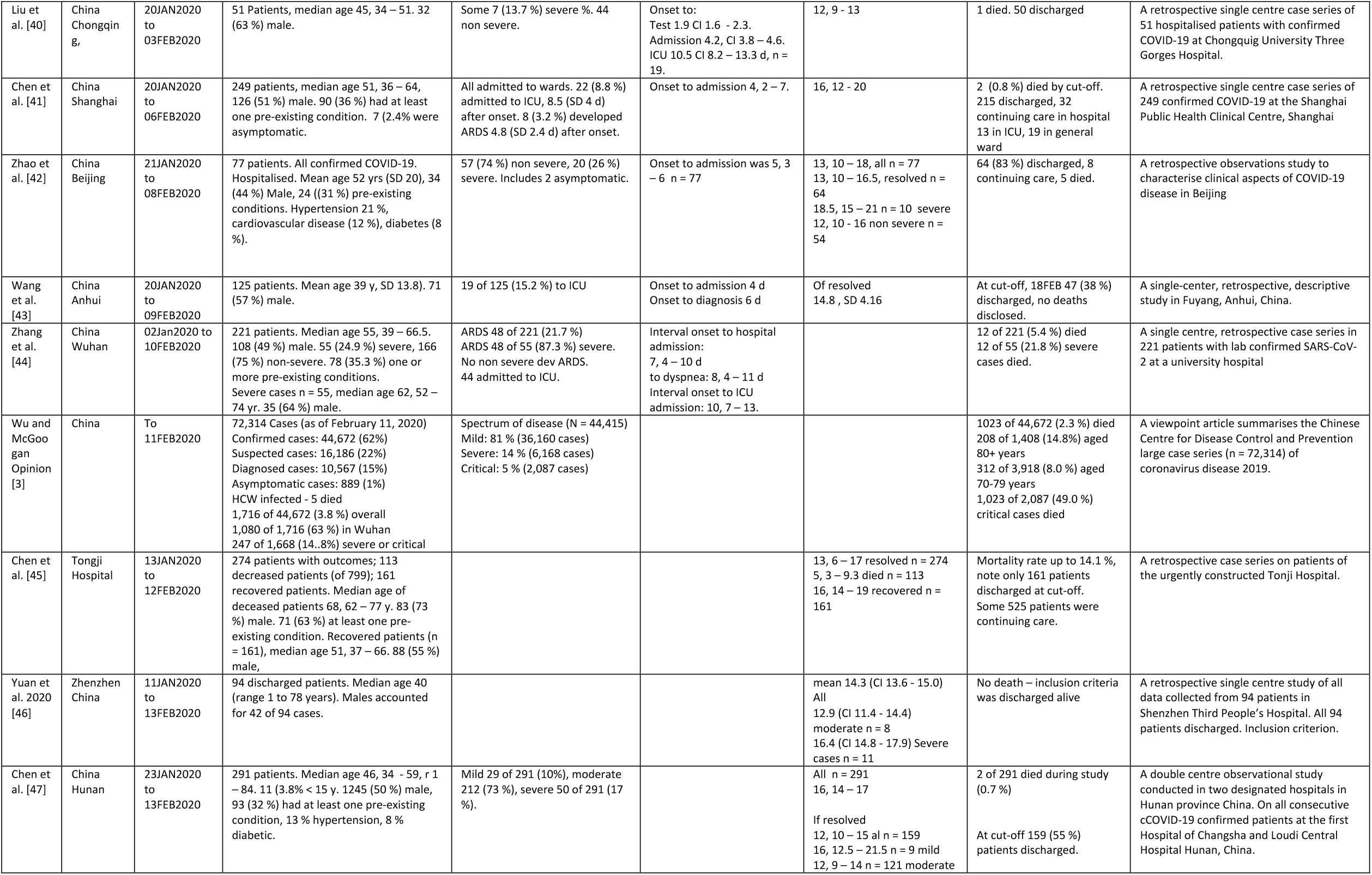
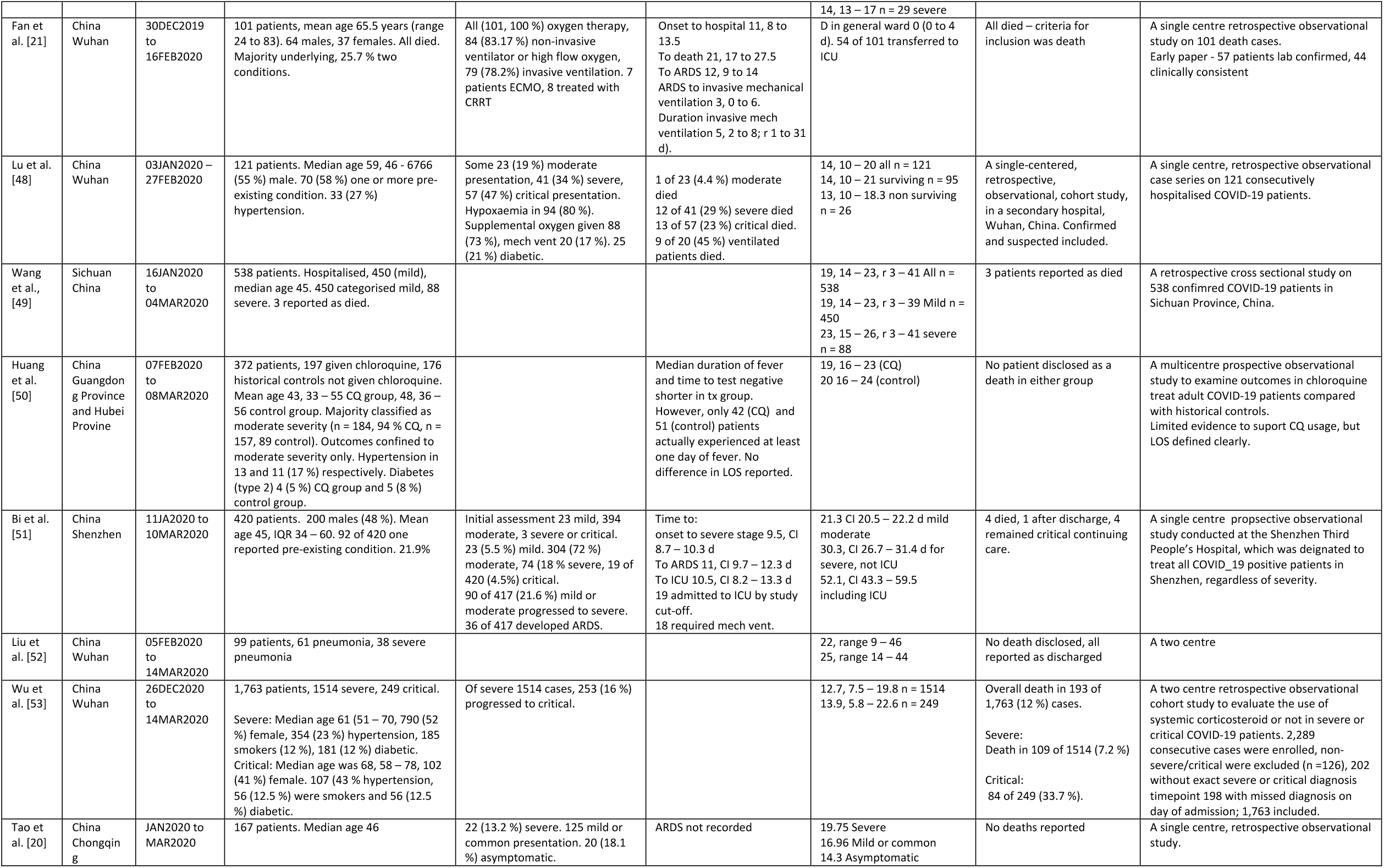
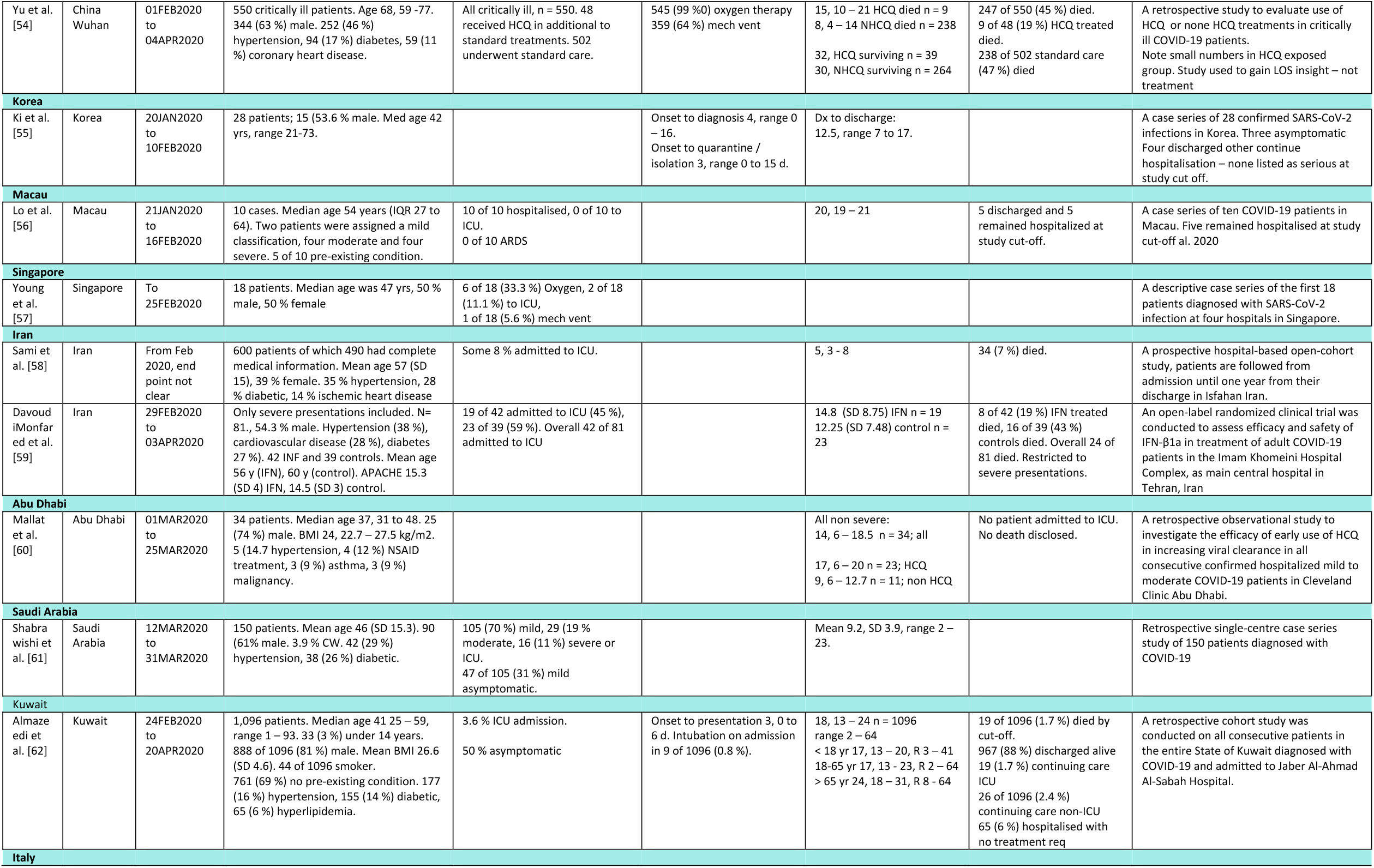
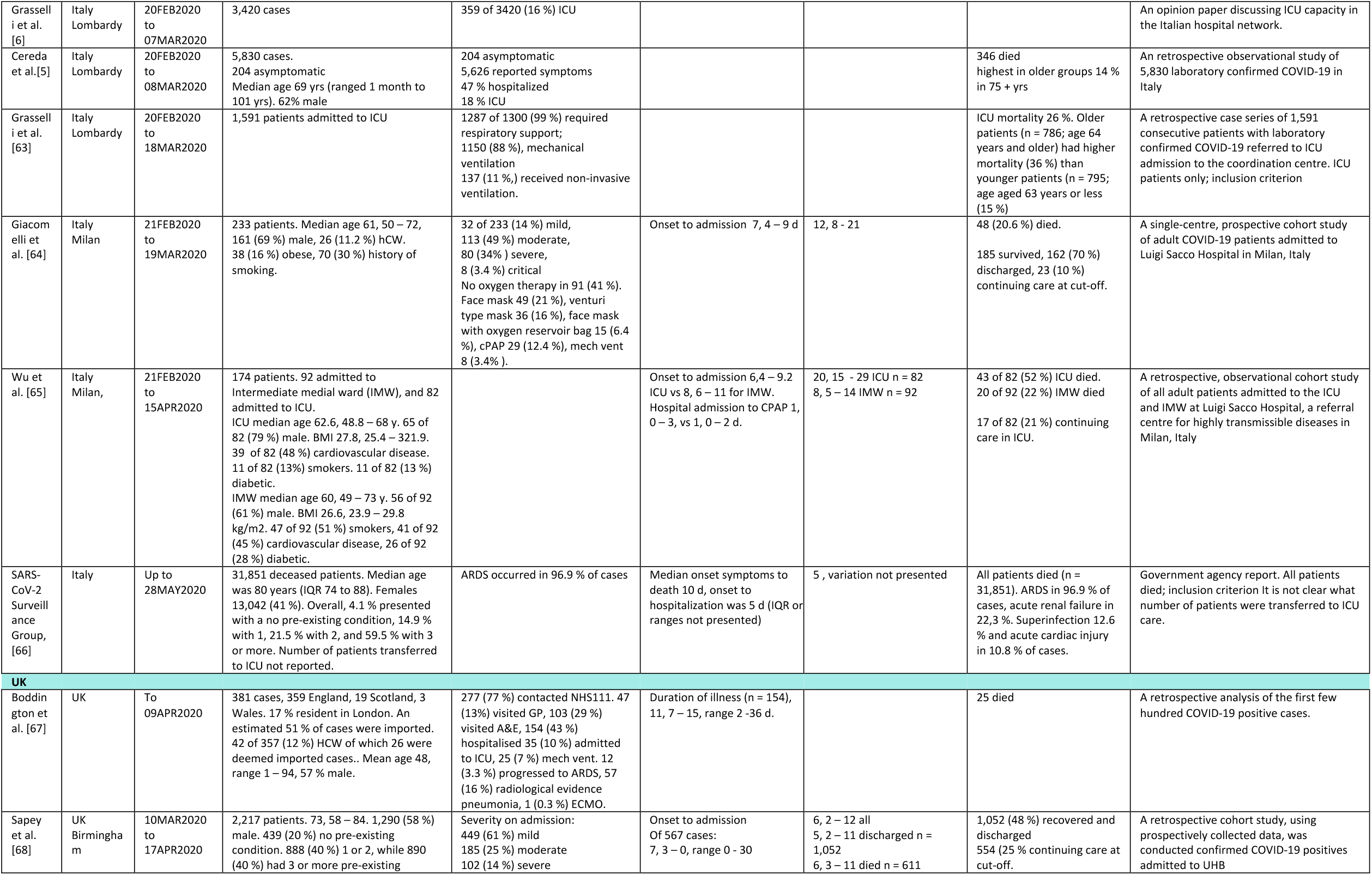
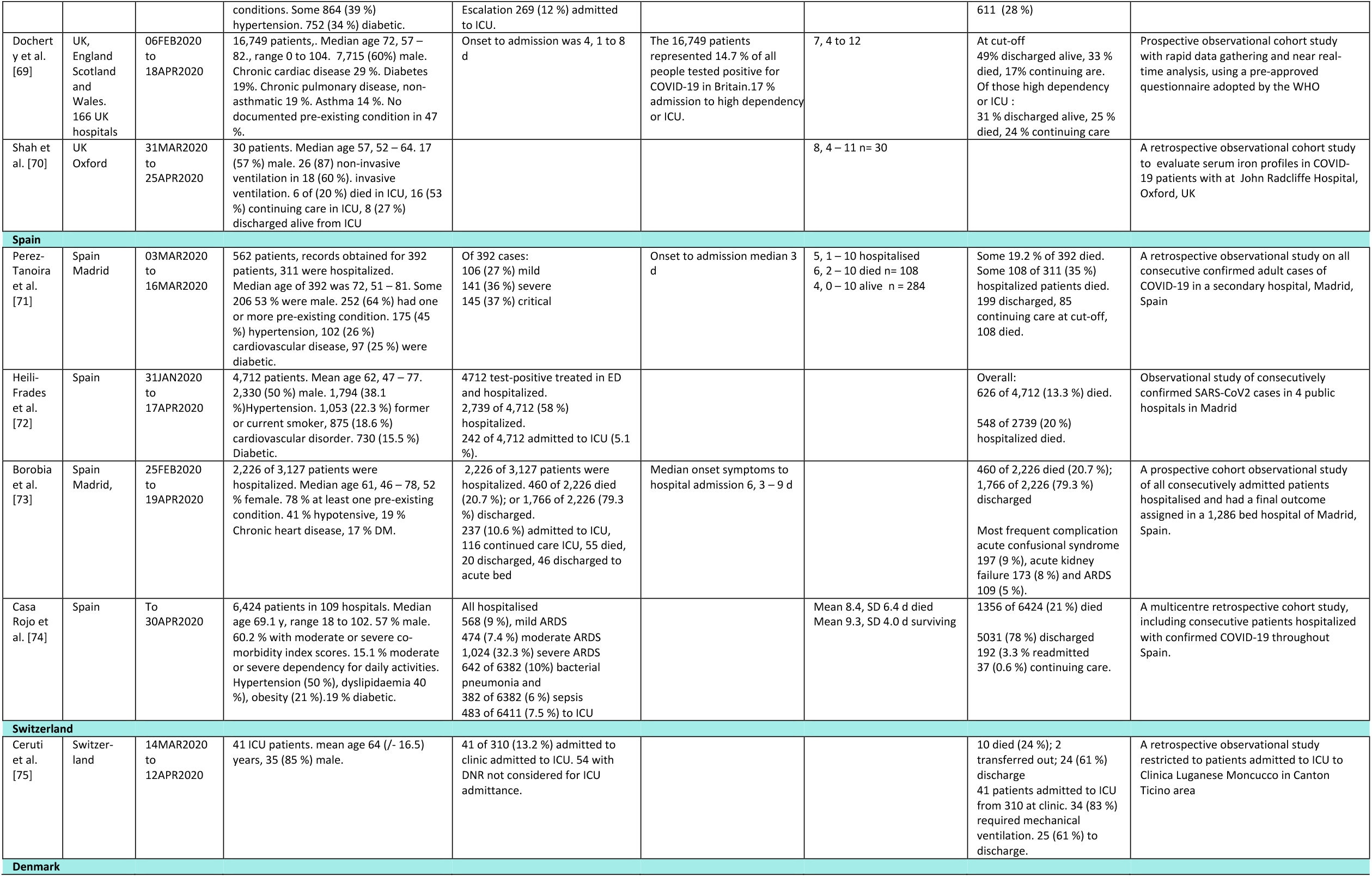
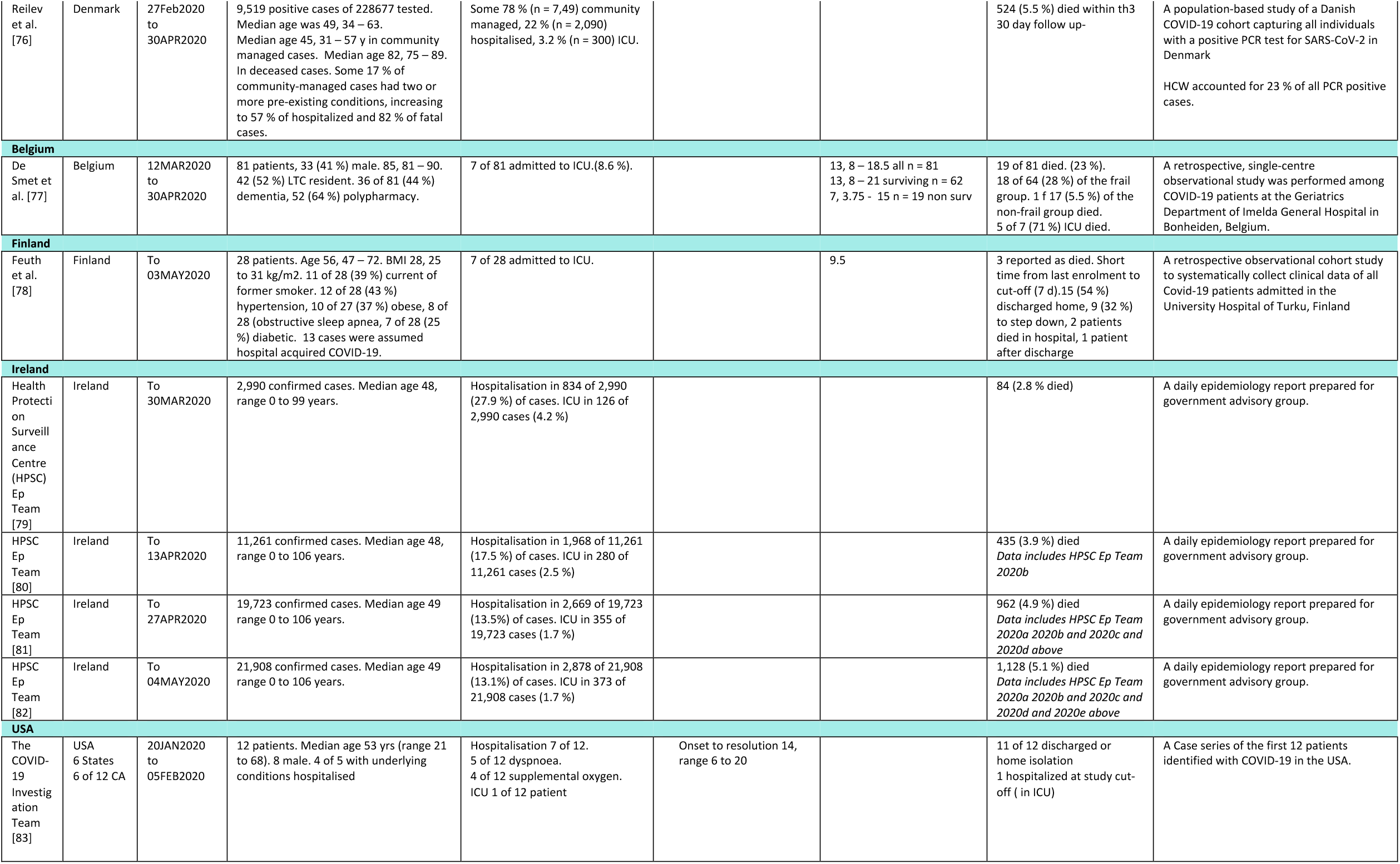
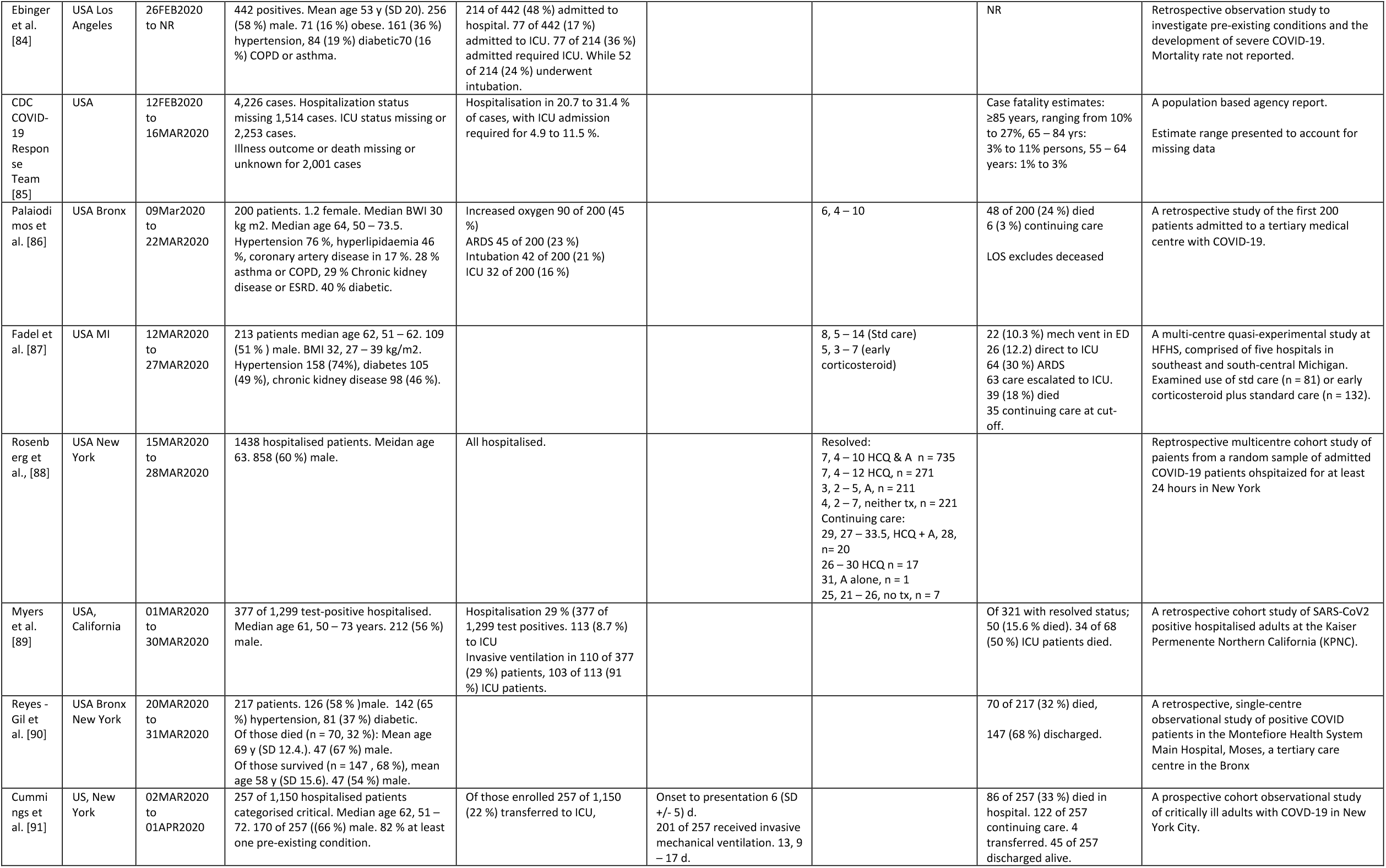
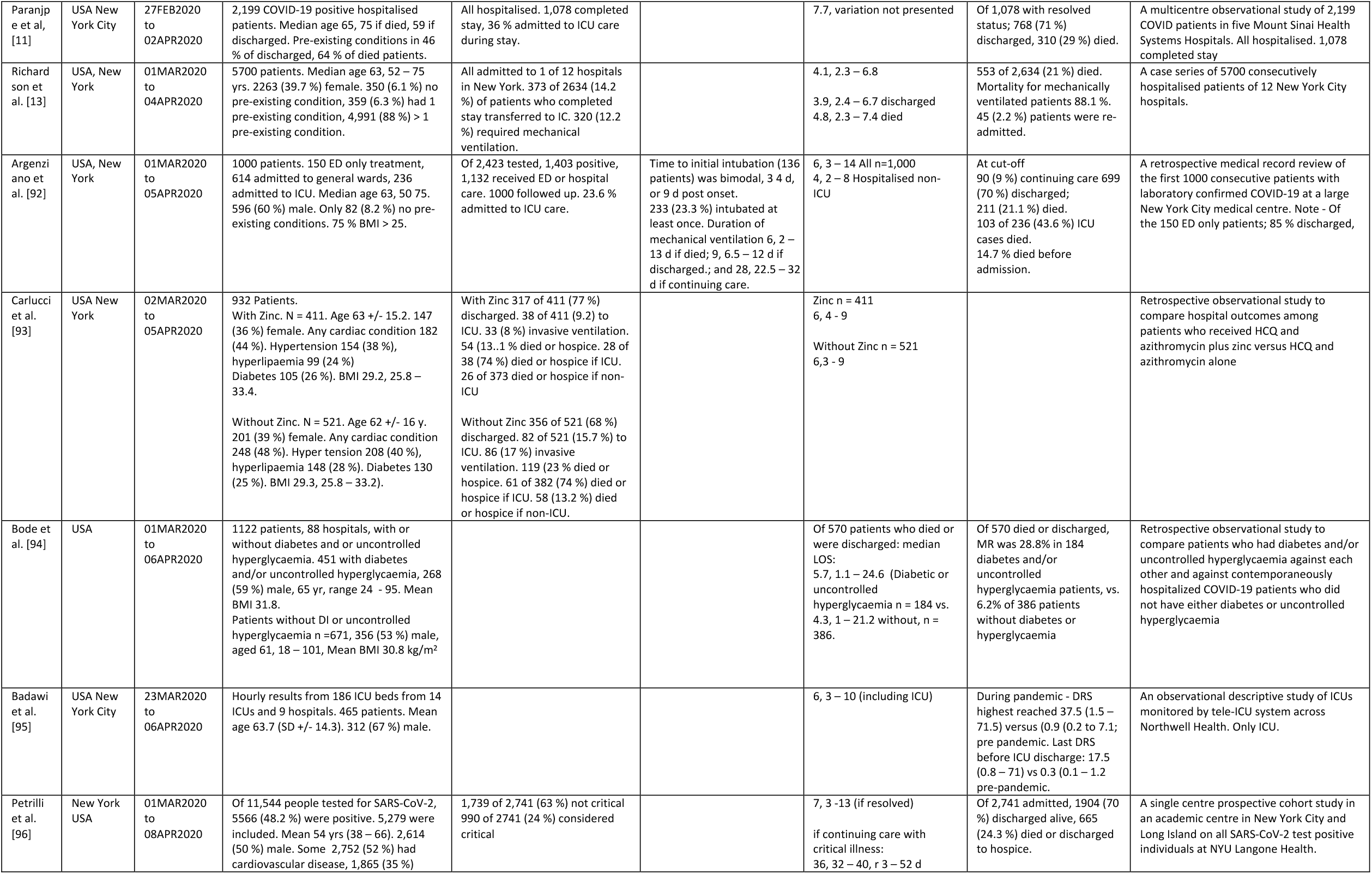
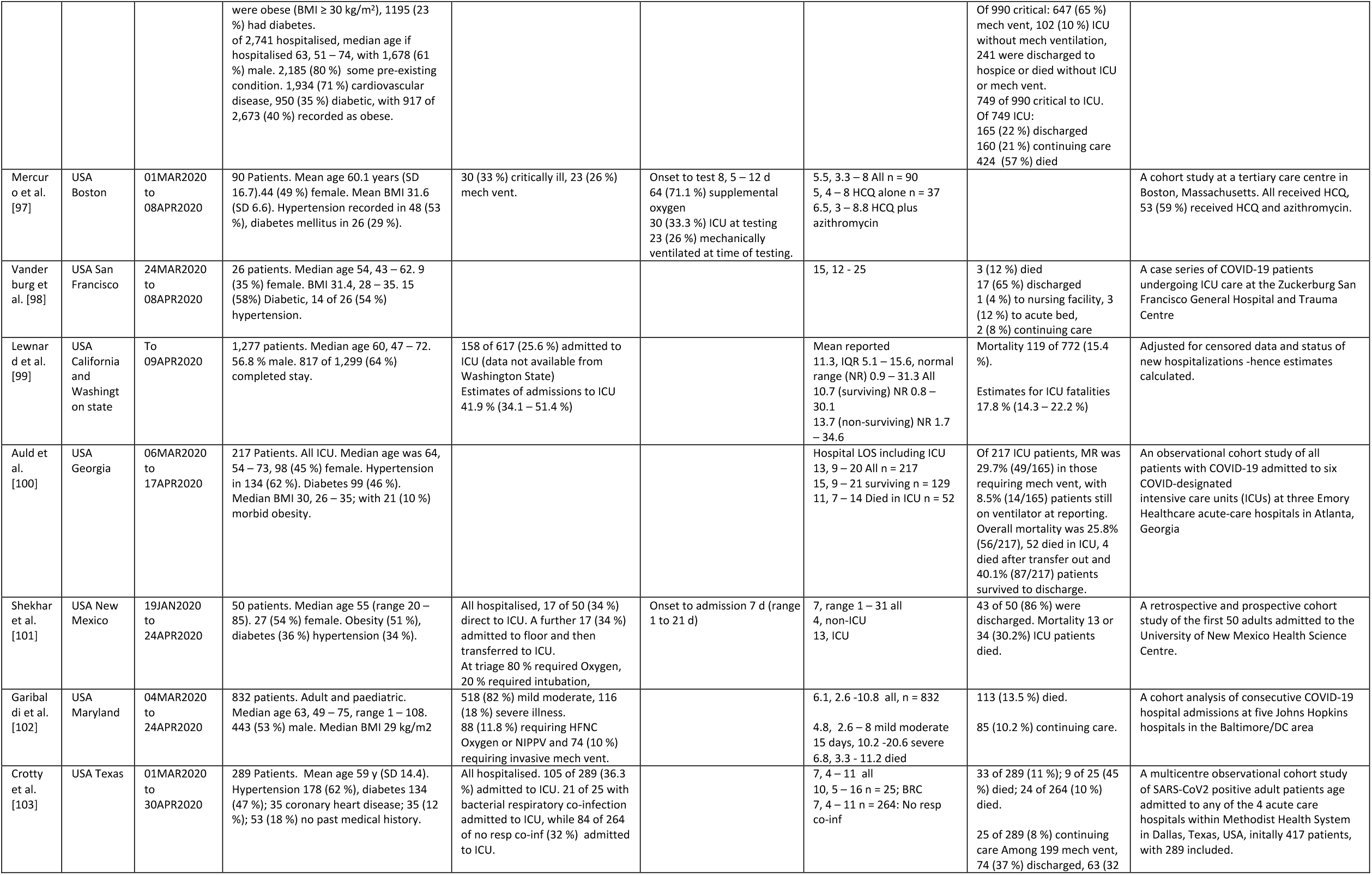
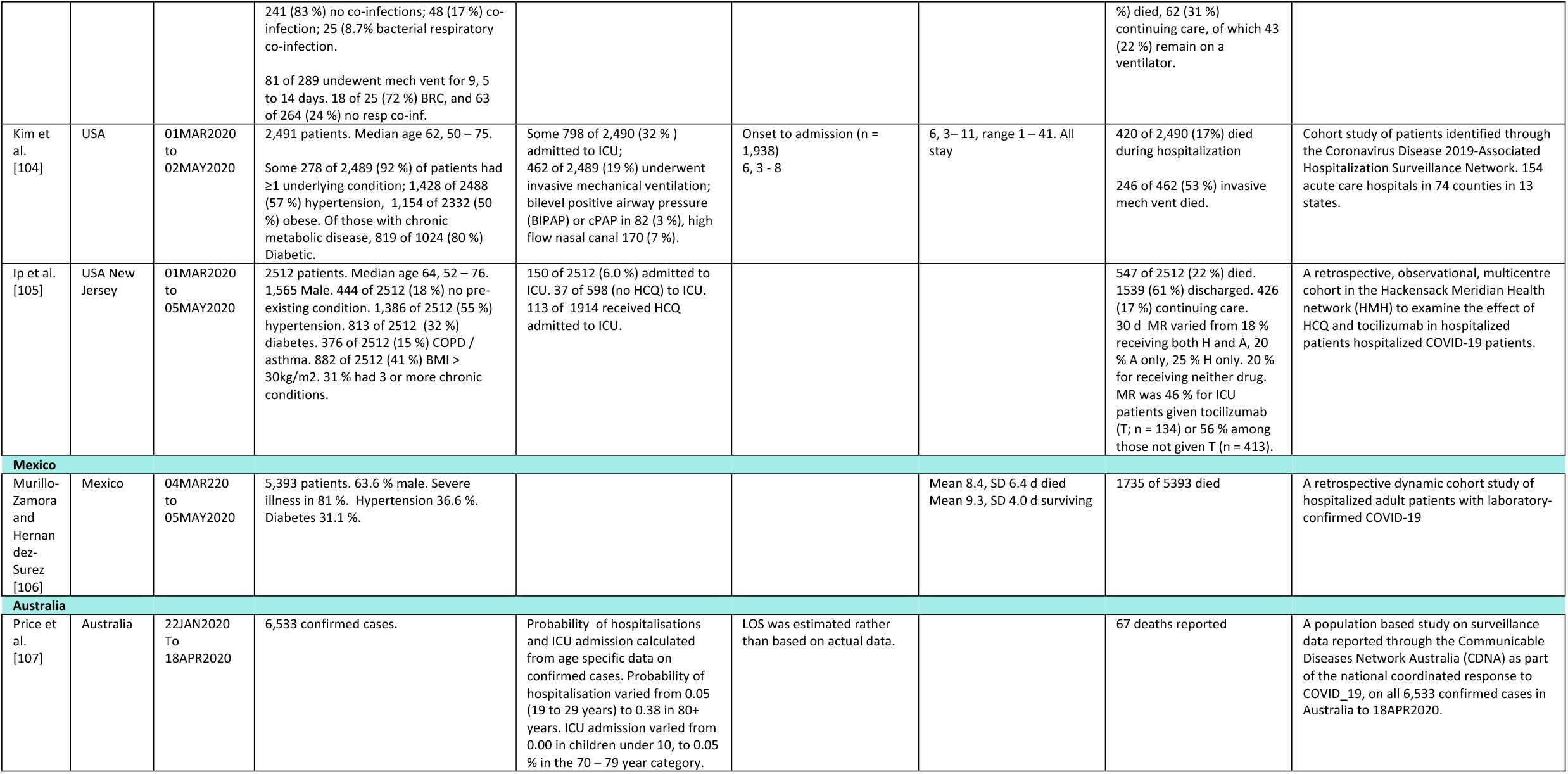
A summary of published articles and reports relating to hospitalisation and length of stay (LOS) in hospital following COVID-19 infection, by country. Ordered by end of enrollment date within country.

#### European Centre for Disease Prevention and Control (ECDC updates)

The European Centre for Disease Prevention and Control (ECDC) collates the laboratory-confirmed cases reported to the European Surveillance System (TESSy) and thus far has published nine update reports as the situation unfolds across Europe [4,7]. Preliminary estimates of severity were collated based on the analysis of data from EU and EEA countries and from the UK. As expected, some data were incomplete or missing in TESSy. The eight update [4] indicates that 48,755 of 152,375 (32 %) were hospitalised based on data from 26 European countries; with a country median 28 % (IQR 14 to 63 %). Severe presentations that required ICU and or respiratory support accounted for 2,859 of 120,788 (2.4 %) cases from 16 countries; median 1.4 % (IQR 0 to 33 %). Among hospitalized cases only, severe illness was reported in 3,567 of 38,960 (9.2 %) cases from 19 countries, median 15 % (IQR 3.8 to 35 %). Death occurred in 1,005 of 9,368 (11 %) of hospitalized cases from 21 countries (3.9 %, IQR 0 to 13 %). Age-specific risk increased among those 60 years and older, thus consideration to age profile within a country is important. The report of 23^rd^ April [7] reported increased rate of hospitalisation with a pooled estimate of 42 % (160,485 of 381,410 cases); based on data from 19 countries, however the country median was 28 % (IQR 16 to 39 %). Severe presentations that required ICU and or respiratory support accounted for 5,456 of 220,412 (2 %) cases with data from 14 countries; median 2 % (IQR 0 to 4 %). Again, hospitalisation increased with age, and for males from 30 years. The changes over time in the crude mortality rate for all notified cases since day of outbreak, defined as the day on which the tenth case was notified, for selected European countries is presented in Figure 2.

**Figure 2.**
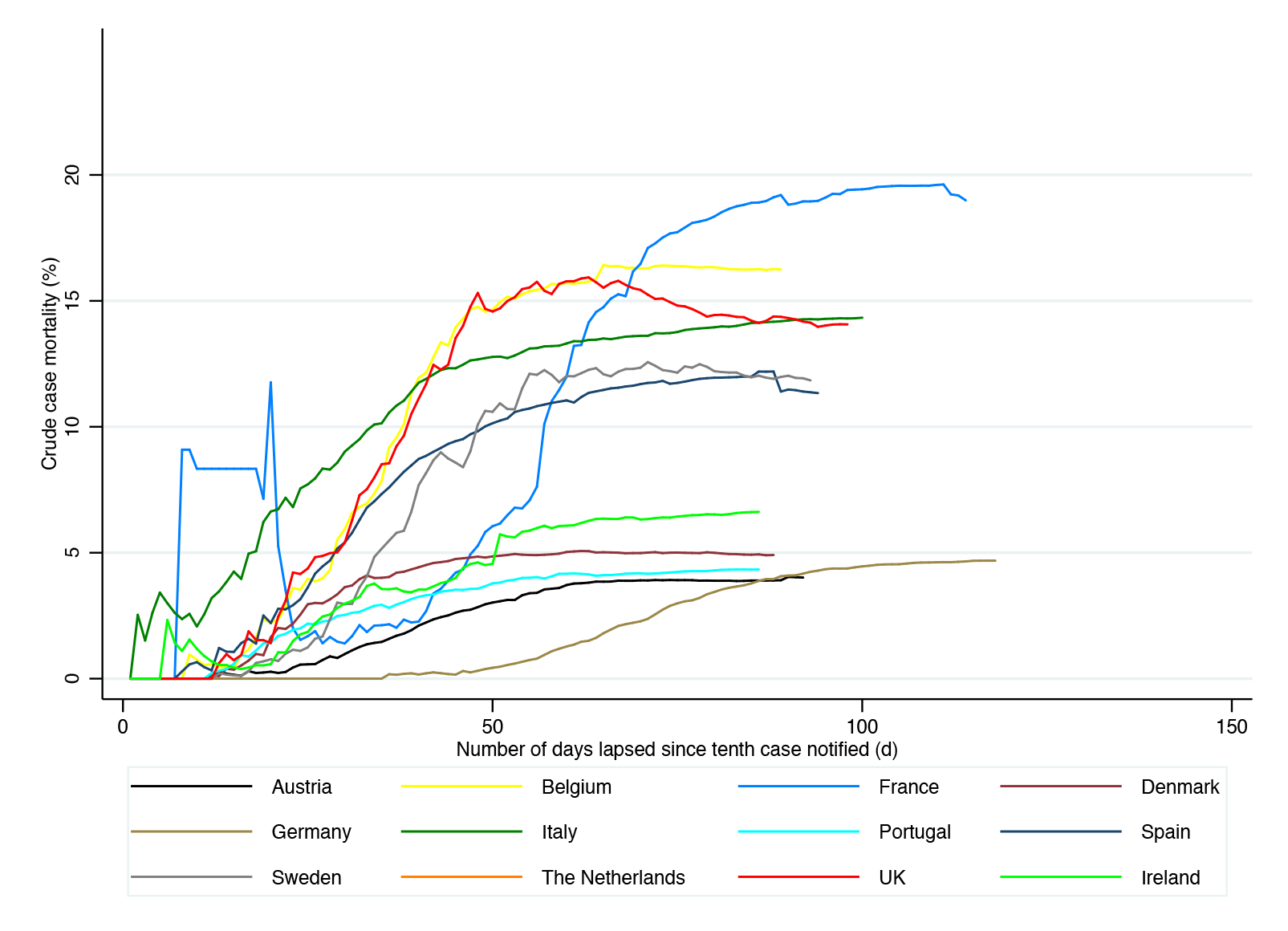
The crude mortality rate (CMR) expressed as the percentage of notified deaths to cases over time, from number of days lapsed since the notification of the tenth case for 12 selected European countries. Data tabulating the notified cases and deaths from all countries is available from the European Centre for Disease Control and Prevention; https://www.ecdc.europa.eu/en/publications-data/download-todays-data-geographic-distribution-covid-19-cases-worldwide

#### USA Studies

Admission rates to ICU in the USA varied between 4.9 to 11.5 % in the report conducted by the CDC [85]. Myers et al. [89] indicated 8.7 % ICU admission in a study conducted in California in March, whereas Lewnard et al. [99] observed a higher ICU admission rate (25.6 %) also in California. A number of studies conducted in New York outlined ICU admission rates of 14.2 % [13]; 22 % [14]; 23.6 % [92]; and 36% [11]. Of significance, are the mortality rates in hospitalised groups (and specifically not ICU only groups) reported in a number of US studies: 15.4 % [89]; 15.6 % [85]; 21 % [13]; 21 % [91]; and 29 % [11].

In Ireland, the proportion of COVID-19 test positives cases (confirmed cases) hospitalised is reducing with time (Figure 3). Daily epidemiology reports compiled by the Health Protection Surveillance Centre [79] with data to March 30^th^ reports a hospitalisation rate of 27.9 % (834 of 2,990 cases hospitalised) and admission to ICU in 4.2 % of cases (126 of 2,990). The hospitalisation rate reduced over time to 17.5 % (1,968 of 11,261) by April 13^th^ [80]; 13.5 % (2,669 of 9,723) by 29^nd^ April [81]; and 13.1 (2,878 of 21,908 cases by 4^th^ May [82]. The rate of admission to ICU reduced, concomitantly, from 4.2 % of cases (126 of 2,990) [79] to 1.7 % (373 of 21,908) [82] of cases by May 4th. The reports collated by the HPSC [33] in Ireland indicate that proportion of hospitalisations and transfer to ICU increased occurred with age.

**Figure 3.**
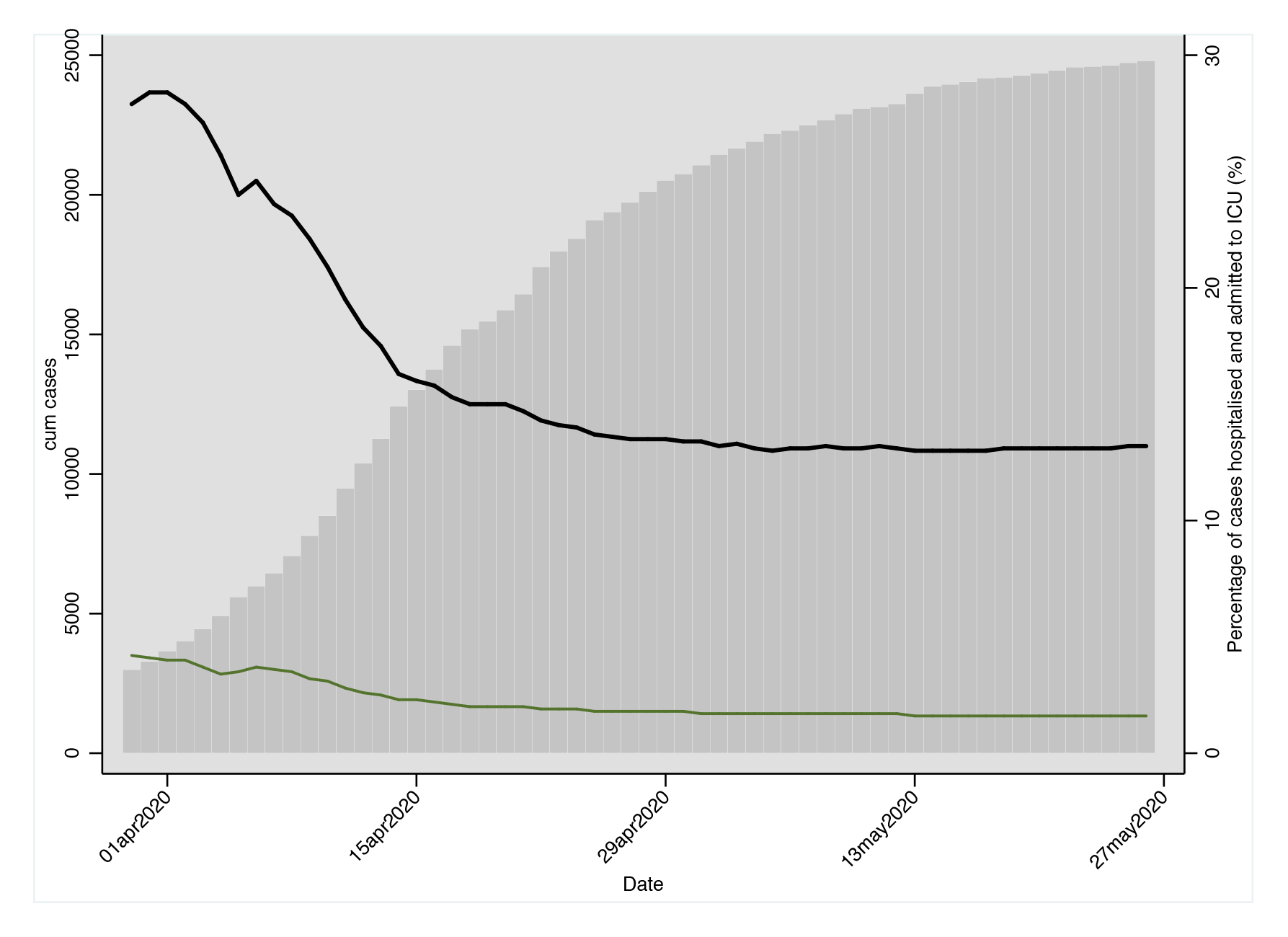
The cumulative number of notified cases (grey bars) in Ireland between 30th March and 27^th^ May 2020; the percentage of COVID positive cases that were hospitalised (black line); and those that were admitted to ICU (green line) for the same period. Data extracted from publicly available data published by the Health Protection Surveillance Centre (HPSC), Ireland.

### LENGTH OF STAY (LOS)

#### Hospital length of stay

A summary of the literature pertaining to length of hospital stay in non-ICU settings due to COVID-19, by country, is reported in Table 1. The overall length of hospital stay is reported in 55 studies or reports, with overall median lengths of stay between three days and fifty-five days. Figure 4 illustrates the median (and IQR or range) of hospital LOS for relevant articles within China (Figure 4a) and outside China (Figure 4b).

**Figure 4.**
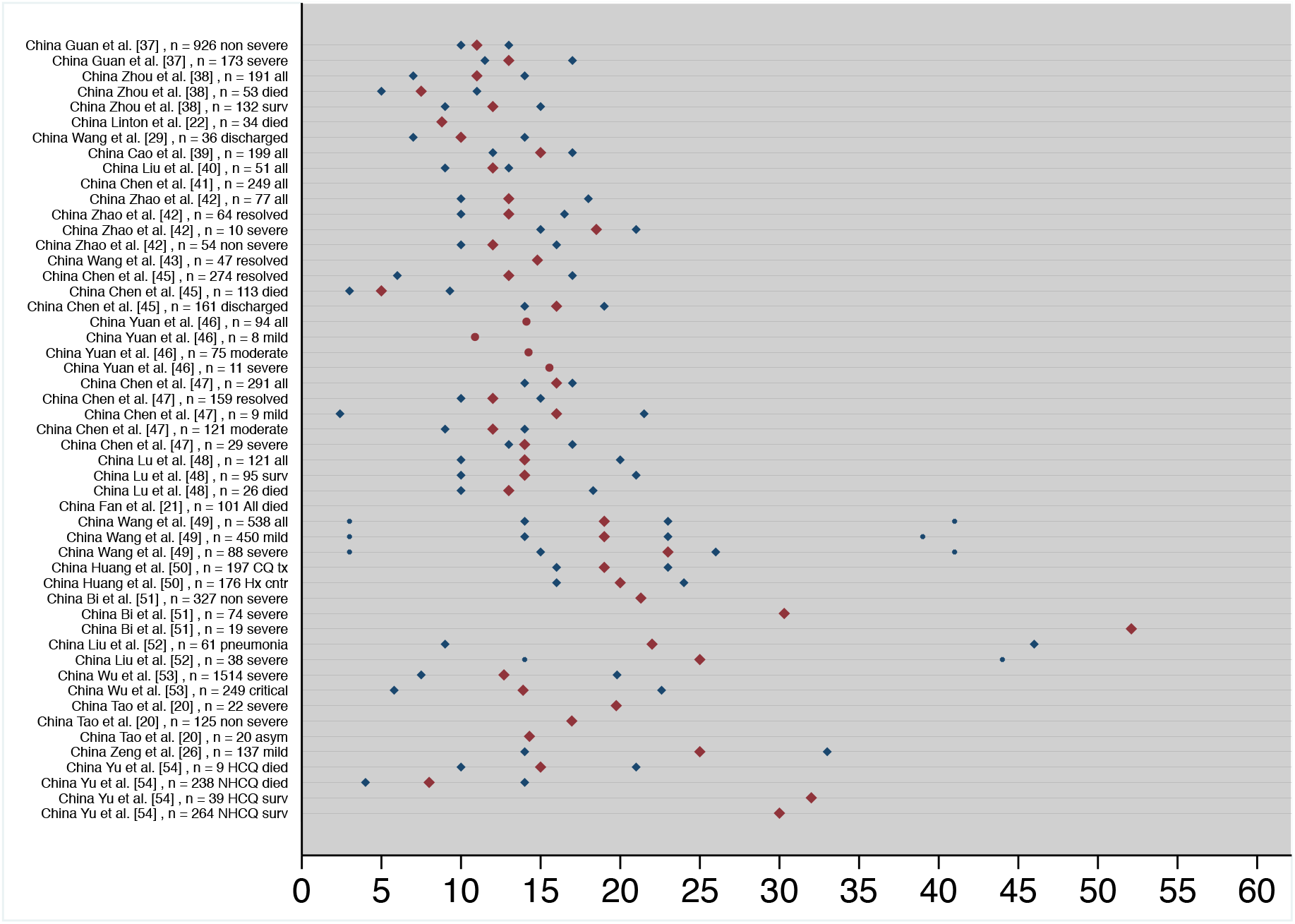
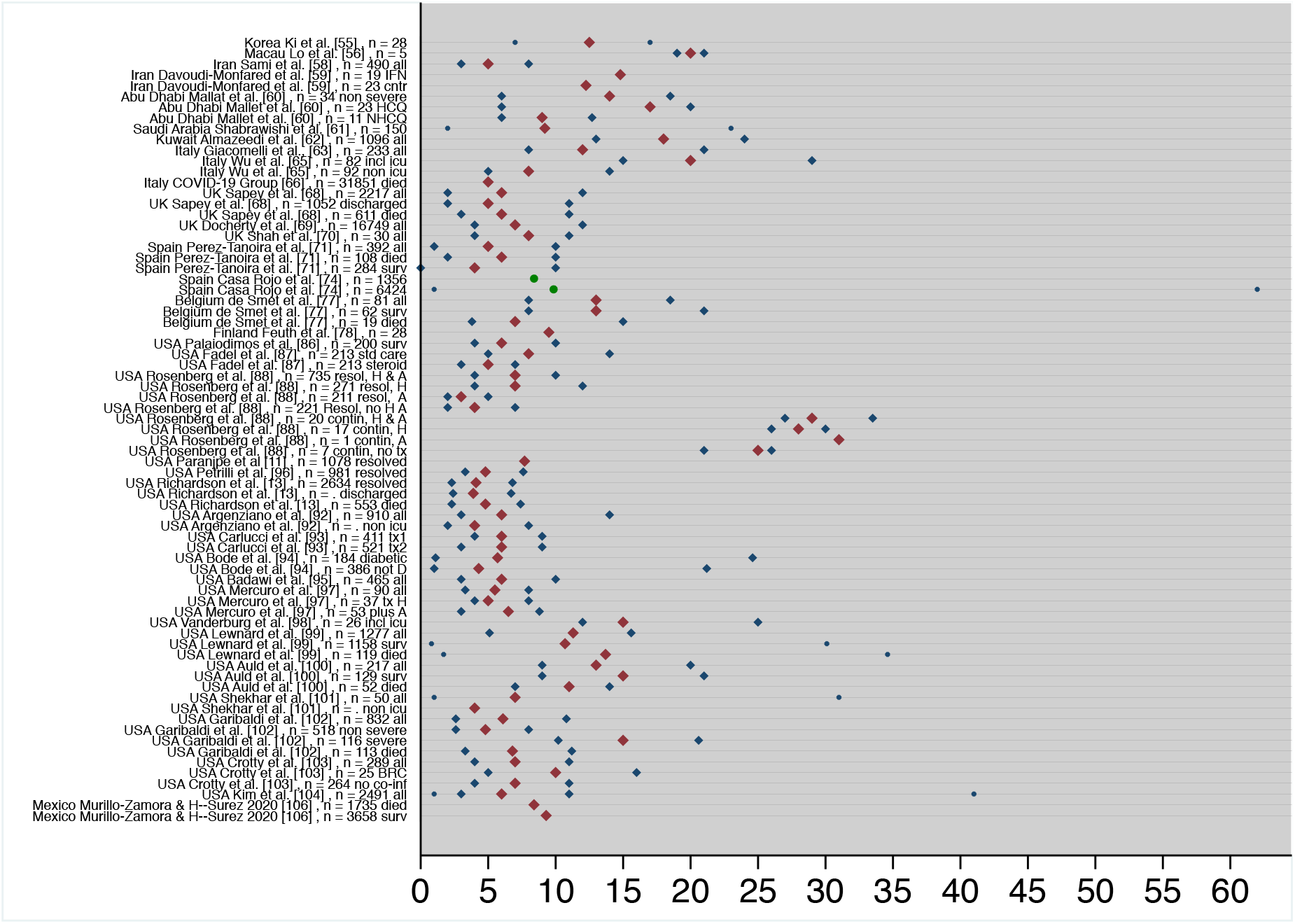
The median (maroon square), or mean (maroon circle), 25^th^ and 75^th^ percentile (navy diamonds) and range (navy x) for relevant articles and reports describing the length of stay (LOS) in hospital due to COVID-19 infection within China (Figure 4a) and outside China (Figure 4b). The Italian report (COVID-19 Group, 2020, n = 31,851) indicates a median of 5 days; no estimate of variance was reported.

#### Length of stay in ICU

The literature pertaining to length of ICU stay due to COVID-19 is reported in Table 2, including 31 studies and reports. Figure 5 illustrates the central tendency (and IQR or range) for length of stay in ICU for relevant articles. In this figure, 26 studies are presented; Three case studies were excluded due to small numbers of relevant cases [8,55,57]. Median lengths of stay in ICU between 7 to 10 days were observed in studies based in China [21,35,38,39,44,51]. European studies conducted by Graselli et al. [63] in Italy, Ceruti et al. [75] in Switzerland, and the ICNARC report [24] report similar LOS between 7 to 11 days. Earlier reports from the ICNARC [23] indicated shorter LOS for ICU patients, indicating the bias towards shorter stay as fewer patients had progress through their complete stay. However median LOS outlined by studies based in North America were more variable and reported lengths of stay between 3.8 to 14 days for various resolved groups (Figure 3); however three studies highlight the continuing care groups with extended median ICU stays of 20 [101], 23 [110] and 36 days [96]. The Reyes-Gil et al. [90] study was excluded from consideration as it was suspected that all patients contributed to the ICU LOS, even if not admitted to ICU.

**Table 2.** A summary of published articles and reports relating to length of stay in an ICU setting following COVID-19 infection, by country.

| Paper | Location | Timing | Demographics | Median LOS pre-ICU (d, IQR) | ICU; LOS median (d, IQR) | Outcomes | Comments |
| --- | --- | --- | --- | --- | --- | --- | --- |
| **China** |  |  |  |  |  |  |  |
| Chen et al. [8] | Wuhan, China | 01JAN2020 to 20JAN2020 | 99 patients. Mean age 55.5 (SD 13.1), 67 male, 32 female. 50 (51 %) chronic conditions. |  | 9 and 11 d ICU n= 2 both died | 11 (11%) died<br>31 (31 %) discharged<br>57 (58 %) still hospitalized by study cut off | A retrospective single centre study including all confirmed COVID19 cases in Wuhan Jinyintan Hospital. |
| Yang et al. [35] | Wuhan, China | DEC 2019 to 26JAN2020 | 52 critically ill patients of 710 in cohort admitted to ICU. Mean age 59.7 (SD 13.3) years, 35 (67 %) male, 21 (40 %) had pre-existing conditions |  | 7, 3–11 ICU to death n= 52 |  | A single centre retrospective observational study that enrolled 52 critically ill patients (ICU only). ICU care only included |
| Zhou et al. [38] | Wuhan, China | To 31JAN2020 | 191 patients. Median age 56 (IQR 46 – 67)<br>Non surviving (median 69 yrs)<br>Surviving (median 52 yrs)<br>Pre-existing 48 % |  | 8, 4 – 12 ICU n = 191<br>7, 2 -9 surviving<br>8, 4 -12 non surviving n= 53 | 54 (28 %) of 191 died<br>132 (72 %) discharged | A retrospective multicentre cohort study of all adult patients at the Jinyintan Hospital and Wuhan Pulmonary Hospital. Median duration of shedding 20 d (longest 37 d) |
| Cao et al. [39] | Wuhan China | 18JAN2020 to 03FEB2020 | 199 patients; age 58, 49 – 68. 120 (60 % male. All severe categorisation, day 1, 28 of 199 did not require supplemental oxygen; category 3), 171 category 4 to 6. Diabetes 23 (12%), cerebrovascular dis 13 (6.5 %), cancer 6 (3 %). |  | 10, 5 – 14 All n = 199<br>10, 8 – 17 surviving<br>10, 4 – 14 non surviving n= 44 | 44 (22.1 %) died | An open-label, individually randomized, controlled trial to evaluate lopinavir – ritonavir treatment and standard care. |
| Zhang et al. [44] | Wuhan, China | 02Jan2020 to 10FEB2020 | 221 patients. Median age 55, 39 – 66.5. 108 (49 %) male. 55 (24.9 %) severe, 166 (75 %) non-severe. 78 (35.3 %) one or more pre-existing conditions. Severe cases n = 55, median age 62, 52 – 74 yr. 35 (64 %) male. |  | 8, 5 – 13 (discharge to ward; n = 23)<br>11 (4.5 to 14.5 (died; n = 9) | 12 of 221 (5.4 %) died<br>12 of 55 (21.8 %) severe cases died. | A single centre, retrospective case series in 221 patients with lab confirmed SARS-CoV-2 at a university |
| Fan et al. [21] | Wuhan China | 30DEC2019 to 16FEB2020 | 101 patients mean age 65.5 years (range 24 to 83). 64 males, 37 females. All died. Majority underlying, 25.7 % two conditions. 47 of 101 direct to ICU: 54 transferred to ICU with aggravating conditions | 0, 0 to 4 n = 54 | 8, 4 to 11. | All died - inclusion criteria | A single centre retrospective observational study on 101 death cases. Early paper - 57 patients lab confirmed, 44 clinically consistent |
| Bi et al. [51] | Shenzhen China | 11JA2020 to 10MAR2020 | 420 patients. 200 males (48 %). Mean age 45, IQR 34 – 60. 92 of 420 one reported pre-existing condition. 21.9 %, 19 admitted to ICU.<br>Onset to ICU admission 10.5, 8.2 – 13.3 d |  | Mean 34.4, CI 24.1 – 43.2 | 4 died, 1 after discharge, 4 remained critical continuing care. All patients were required to be hospitalized for about two weeks for isolation. Mech vent 28.5, CI 20 – 39.1 d | A single centre prospective observational study conducted at the Shenzhen Third People's Hospital, which was designated to treat all COVID-19 positive patients in Shenzhen, regardless of severity. |
| **Singapore** |  |  |  |  |  |  |  |
| Young et al. [57] | Singapore | To 25FEB2020 | 2 of 18 admitted to ICU. Median age was 47 yrs, 50 % male, 50 % female | Mean 6 (range 5 – 7) | Mean 6 (range 5 – 7) |  | A descriptive case series of the first 18 patients diagnosed with SARS-CoV-2 infection at four hospitals in Singapore. |
| **Iran** |  |  |  |  |  |  |  |
| Davoudi-Monfared et al. [59] | Iran | 29FEB2020 to 03APR2020 | Only severe presentations included. N= 81., 54.3 % male. Hypertension (38 %), cardiovascular disease (28 %), diabetes 27 %). 42 INF and 39 controls. Mean age 56 y (INF), 60 y (control). APACHE 15.3 (SD 4) INF, 14.5 (SD 3) control. |  | 7.71 (SD 8.75) INF<br>8.25 (SD 7.48) control | 8 of 42 (19 %) INF treated died, 16 of 39 (43 %) controls died. Overall 24 of 81 died. Restricted to severe presentations. | An open-label randomized clinical trial was conducted to assess efficacy and safety of IFN-β1a in treatment of adult COVID-19 patients in the Imam |
|  |  |  | 42 of 81 admitted to ICU. 19 of 42 IFN admitted to ICU (45 %), 23 of 39 (control; 59 %). Overall 42 of 81 admitted to ICU. |  |  |  | Khomeini Hospital Complex, as main central hospital in Tehran, Iran |
| **Italy** |  |  |  |  |  |  |  |
| Grasselli et al. [63] | Lombardy Italy | 20FEB2020 to 18MAR2020 | 1,591 critically ill patients admitted to ICU, with 1,581 ICU info available.<br>Of 1043 pre-existing conditions available: 334 (32 %) none, 509 (49 %) hypertension, 223 (21 %) cardiovascular disease; 188 (18 %) hypercholesterolaemia; 180 (17 %) diabetic.<br>Of 130 with resp support data: 1150 (88%) required IMV 137 (11) non invasive MV 13 (1 %) Oxygen mask |  | 9, 6 – 13; All 10, 8 – 14; continuing care n = 920<br>8, 5 – 12; discharged n = 256<br>7, 5 – 11; died n = 405 | ICU mortality 26 %.<br>64 + yrs (n = 786) 36 % died<br>63 or younger (n = 795) 15 % died.<br>At study cut off: 405 of 1,581 (26 %) died in ICU 256 of 1,591 (16 %) discharged 920 of 1,591 (58 %) continued in ICU. | A retrospective case series of 1,591 consecutive patients with laboratory confirmed COVID-19 referred to ICU admission to the coordination centre. ICU patients only; inclusion criterion |
| Wu et al. [108] | Milan, Italy | 21FEB2020 to 15APR2020 | 174 patients. 92 admitted to Intermediate medial ward (IMW), and 82 admitted to ICU.<br>ICU 62.6, 48.8 – 68. 65 of 82 (79 %) male. BMI 27.8, 25.4 – 321.9. 39 of 82 (48 %) cardiovascular disease. 11 of 82 (13%) smokers. 11 of 82 (13 %) diabetic.<br>IMW 60, 49 – 73. 56 of 92 (61 %) male. BMI 26.6, 23.9 – 29.8 kg/m2. 47 of 92 (51 %) smokers, 41 of 92 (45 %) cardiovascular disease, 26 of 92 (28 %) diabetic. |  | 13, 7.7 – 17 resolved | 43 of 82 (52 %) ICU died.<br>20 of 92 (22 %) IMW died<br>17 of 82 (21 %) continuing care<br>3 of 82 (3.7 %) continuing care in ICU. | A retrospective, observational cohort study of all adult patients admitted to the ICU and IMW at Luigi Sacco Hospital, a referral centre for highly transmissible diseases in Milan, Italy |
| SARS-CoV-2 Surveillance Group [66] | Italy | Up to 28MAY2020 | 31,851 deceased patients. Median age was 80 years (IQR 74 to 88). Females 13,042 (41 %). Overall, 4.1 % presented with a no pre-existing condition, 14.9 % with 1, 21.5 % with 2, and 59.5 % with 3 or more. Number of patients transferred to ICU not reported. |  | 9 (if included ICU care; IQR or range not presented). | All patients died (n = 31,851). ARDS in 96.9 % of cases, acute renal failure in 22.3 %. Superinfection 12.6 % and acute cardiac injury in 10.8 % of cases. | All patients died; inclusion criterion It is not clear what number of patients were transferred to ICU care. |
| **Switzerland** |  |  |  |  |  |  |  |
| Ceruti et al. [75] | Switzerland | 14MAR2020 to 12APR2020 | 41 ICU patients. mean age 64 (/– 16.5) years, 35 (85 %) male. 19 (46 %) hypertension; 15 (37 %) diabetic.<br>41 of 310 to ICU.<br>Of ICU (n = 41) 34 (83 %) intubated, 7 (17 %) non intubated. |  | 9, 4 – 12.5; discharged | 10 died (24 %); 2 transferred out; 24 (61 %) discharge. 4 (10 %) continuing care (MV)<br>41 patients admitted to ICU from 310 at clinic. 34 (83 %) required mechanical ventilation. 25 (61 %) to discharge. Median duration of MV 7, 3 – 10 d. | A retrospective observational study restricted to patients admitted to ICU to Clinica Luganese Moncucco in Canton Ticino area |
| **Finland** |  |  |  |  |  |  |  |
| Feuth et al. [78] | Finland | To 03MAY2020 | 28 patients. Age 56, 47 – 72. BMI 28, 25 to 31 kg/m2. 11 of 28 (39 %) current of former smoker. 12 of 28 (43 %) hypertension, 10 of 27 (37 %) obese, 8 of 28 (obstructive sleep apnea, 7 of 28 (25 %) diabetic. 7 of 28 (25 %) admitted to ICU. |  | 19 | 3 reported as died. Short time from last enrolment to cut-off (7 d). 2 continuing care (1 in ICU) 15 (54 %) discharged home, 9 (32 %) to step down, 2 | A retrospective observational cohort study to systematically collect clinical data of all Covid-19 patients admitted in the University Hospital of Turku, Finland |
|  |  |  |  |  |  | patients died in hospital, 1 patient after discharge to step down care. |  |
| **UK** |  |  |  |  |  |  |  |
| ICNARC, [24]<br>29MAY2020 | UK | Up to<br>29MAY2020 | 9,347 Patients. Age 60, 51 – 68. 6,621 (71 %) Male. 6,389 of 8,638 (74 %) patients had BMI ≥ 25. | 1, 0 to 3 Pre ICU LOS, n = 8,233 | 11, 4 – 22; Surviving n = 4,579<br>9, 5 - 15; Non surviving, n = 3,483<br>1, 0 to 3 Pre ICU LOS, n = 7,524 | 8,062 of 9,347 completed ICU care:<br>4,579 of 8,062 (57 %) alive<br>3,483 of 8,062 (43 %) died | Government agency report. 5,437 of 8,818 (62 % mech vent within 24 hrs APACHE II score 14, 11 – 18; n = 9,008 |
| Shah et al. [70] | UK Oxford | 31MAR2020 to<br>25APR2020 | 30 patients. Median age 57, 52 – 64. 17 (57 %) male. All critically ill COVID-19 in ICU.<br>26 (87) invasive ventilation, 18 (60 %) non-invasive ventilation. |  | 8, 4 – 11, n= 30 | 6 of 30 (20 %) died in ICU, 16 of 30 (53 %) continuing care in ICU, 8 of 30 (27 %) discharged alive from ICU | A retrospective observational cohort study to evaluate serum iron profiles in COVID-19 patients with at John Radcliffe Hospital, Oxford, UK |
| **USA** |  |  |  |  |  |  |  |
| The COVID-19 Investigation Team et al. [83] | USA<br>6 States<br>6 of 12 CA | 20JAN2020 to<br>05FEB2020 | 12 patients. Median age 53 yrs (range 21 to 68). 8 male. 4 of 5 with underlying conditions hospitalised. First case series in USA. Only 1 admitted to ICU. | 5 d | 5 | 11 of 12 discharged or remained home isolation<br>1 remained hospitalized at study cut off (patient who was admitted to ICU) | A Case series of the first 12 patients identified with COVID-19 in the USA. |
| Bhatraju et al. [109] | Seattle<br>USA | 24FEB2020 to<br>09MAR2020 | 24 Patients. Mean age 64 ± 18 yrs, 63 % male. Critically ill only and all admitted to an ICU. 14 (58 %) diabetes; 5 (21 %) chronic kidney disease. 8 (33 %) had two or more pre-existing condition. 5 (22 %) current or former smoker; 5 (21 %) obstructive sleep apnea.<br>Onset to admission 7 (SD 4 d). |  | 9, 4 to 14 All<br>14, 4 – 17 surviving only | 12 of 24 died<br>Surviving only:<br>5 discharged home,<br>4 discharged to general wards,<br>3 remained on mechanical ventilation | A multicentre observational study including COVID-19 patients admitted to one of nine hospital ICUs. Only ICU patients. ICU, inclusion criterion |
| Reyes- Gil et al. [90] | USA Bronx<br>New York | 20MAR2020 to<br>31MAR2020 | 217 patients. 126 (58 %) male. 142 (65 %) hypertension, 81 (37 %) diabetic.<br>If died (n = 70, 32 %): Mean age 69 y (SD 12.4). 47 (67 %) male.<br>Survivors (n = 147, 68 %), mean age 58 y (SD 15.6). 47 (54 %) male. |  | 0, 0 – 0 died n = 70<br>0, 0 – 5<br>Misreported – included those not placed in ICU. | 70 of 217 (32 %) died,<br>147 (68 %) discharged. | A retrospective, single-centre observational study |
| Cummings et al. [91] | New York,<br>USA | 02MAR2020 to<br>01APR2020 | 257 of 1,150 hospitalised patients categorised critical. Median age 62, 51 – 72. 170 of 257 (66 %) male. 82 % at least one pre-existing condition. |  | 8, 5 – 12 (died only) | 86 of 257 (33 %) died in hospital. 122 of 257 continuing care. 4 transferred. 45 of 257 discharged alive. 201 of 257 received invasive mechanical ventilation. 13, 9 – 17 d. | A prospective cohort observational study of critically ill adults with COVID-19 in New York City. |
| Argenziano et al. [92] | New York,<br>USA | 01MAR2020 to<br>05APR2020 | 1000 patients. 150 ED only treatment, 614 admitted to general wards, 236 to ICU. Median age (all) 63, 50-75. 596 (60 %) male. Only 82 (8.2 %) no pre-existing conditions. 75 % BMI > 25.2. Median age (ICU n = 236) 62, 52 – 72. 158 of 236 (67 %) male. 75 % BMI > 25.7.<br>ARDS 35 % of all, and 90 % ICU patients. 220 (93 %) intubated at least once, 74 (31 %) extubated at least once. |  | 23, 12 – 32 includes ICU | At cut-off in ICU:<br>87 (37 %) continuing care<br>46 (20 %) discharged;<br>103 (44 %) ICU cases died<br>Of the 150 ED only patients;<br>85 % discharged, 14.7 % died before admission.<br>Median time intubated 6, 2 – 13 died, n = 107; 9, 6.5 -12 | A retrospective medical record review of the first 1000 consecutive patients with laboratory confirmed COVID-19 at a large New York City medical centre. LOS include hospital stay and also includes dead, discharged and continuing care. |
|  |  |  |  |  |  | discharged, n = 31; 28.5, 22.5 – 31.75) continuing care, n = 86. |  |
| Carlucci et al. [93] | New York, USA | 02MAR2020 to 05APR2020 | 932 Patients.<br>With Zinc. N = 411. Age 63 +/- 15.2. 147 (36 %) female. Any cardiac condition 182 (44 %). Hypertension 154 (38 %), hyperlipaemia 99 (24 %), diabetes 105 (26 %). BMI 29.2, 25.8 – 33.4.<br>Without Zinc. N = 521. Age 62 +/- 16 y. 201 (39 %) female. Any cardiac condition 248 (48 %). Hyper tension 208 (40 %), hyperlipaemia 148 (28 %). Diabetes 130 (25 %). BMI 29.3, 25.8 – 33.2). |  | Zinc n = 38<br>4.85, 1.97 – 7.94<br>Without Zinc n = 82<br>5.54, 2.65 – 9.32 | With Zinc 317 of 411 (77 %) discharged. 54 (13.1 %) died or hospice. 28 of 38 (74 %) died or hospice if ICU. 26 of 373 died or hospice if non-ICU<br>Without Zinc 356 of 521 (68 %) discharged. 119 (23 %) died or hospice. 61 of 382 (74 %) died or hospice if ICU. 58 (13.2 %) died or hospice if non-ICU. | Retrospective observational study to compare hospital outcomes among patients who received HCQ and azithromycin plus zinc versus HCQ and azithromycin alone |
| Badawi et al. [95] | USA New York City | 23MAR2020 to 06APR2020 | Data relates to 465 patients. Mean age 63.7 (SD +/- 14.3). 312 (67 %) male. |  | 3.8, 1.4 – 7.3<br>4.2, 1.8 – 7.7 (Mechanically ventilated) |  | Study examined ICU occupancy. During pandemic - DRS highest reached 37.5 (1.5 – 71.5) versus (0.9 (0.2 to 7.1; pre pandemic. Last DRS before ICU discharge: 17.5 (0.8 – 71) vs 0.3 (0.1 – 1.2 pre-pandemic. |
| Petrilli et al. [96] | New York USA | 01MAR2020 to 08APR2020 | Of 11,544 people tested for SARS-CoV-2, 5566 (48.2 %) were positive. 5,279 were included. Mean 54 yrs (38 – 66). 2,614 (50 %) male. Some 2,752 (52 %) had cardiovascular disease, 1,865 (35 %) were obese (BMI ≥ 30 kg/m<sup>2</sup>), 1195 (23 %) had diabetes.<br>Some 2,741 were hospitalised, median age if hospitalised 63, 51 – 74, with 1,678 (61 %) male. 2,185 (80 %) some pre-existing condition. 1,934 (71 %) cardiovascular disease, 950 (35 %) diabetic, with 917 of 2,673 (40 %) recorded as obese.<br>1,739 of 2,741 (63 %) not critical<br>990 of 2741 (24 %) considered critical |  | Critical illness n = 990 (includes hospital stay)<br>9, 5 – 17 if discharged or died<br>if ongoing (n = 160)<br>36, 32 – 40, r 3 – 52 d | Of 990 critical: 647 (65 %) mech vent, 102 (10 %) ICU without mech ventilation, 241 were discharged to hospice or died without ICU or mech vent.<br>749 of 990 critical to ICU.<br>Of 749 ICU:<br>165 (22 %) discharged<br>160 (21 %) continuing care<br>424 (57 %) died or hospice care | A single centre prospective cohort study in an academic centre in New York City and Long Island on all SARS-CoV-2 test positive individuals at NYU Langone Health. |
| Vanderburg et al. [98] | USA San Francisco | 24MAR2020 to 08APR2020 | 26 patients. Median age 54, 43 – 62. 9 (35 %) female. BMI 31.4, 28 – 35. 15 (58%) Diabetic, 14 of 26 (54 %) hypertension. All ICU. 20 of 26 (77 %) mechanically vent at some point. Duration of MV 13.5, 5 – 20 d.<br>11 (42 %) direct to ICU, 15 (58 %) stepped up care | 2, 1 – 2, n = 15 | 9, 3 - 24 | 3 (12 %) died<br>17 (65 %) discharged home,<br>1 (4 %) to nursing facility, 3 (12 %) to acute bed,<br>2 (8 %) continuing care | A case series of COVID-19 patients undergoing ICU care at the Zuckerberg San Francisco General Hospital and Trauma Centre |
| Lewnard et al. [99] | USA California and Washington state | To 09APR2020 | 1,277 patients. Median age 60, 47 – 72. 56.8 % male. 817 of 1,299 (64 %) completed stay. 158 of 617 (25.6 %) admitted to ICU (data not available from Washington State) |  | Mean, IQR<br>8.1, 3.9 – 11.2 | Estimates of admissions to ICU<br>41.9 % (34.1 – 51.4 %)<br>Estimates for ICU fatalities<br>17.8 % (14.3 – 22.2 %) | A multicentre prospective cohort observational study on patients admitted to ICU care due to COVID-19. Adjusted for censored data hence estimates calculated. |
| Auld et al. [100] | USA Georgia | 06MAR2020 to 17APR2020 | 217 Patients. All ICU. Median age was 64, 54 – 73, 98 (45 %) female. Hypertension in 134 (62 %). Diabetes 99 (46 %). Median BMI 30, 26 – 35; with 21 (10 %) morbid obesity. |  | 8, 3 – 13 All n = 217<br>7, 3 – 12 surviving = 129<br>8, 5 – 12 died, n = 52 | Of 217 ICU patients, MR was 29.7% (49/165) in those requiring mech vent, with 8.5% (14/165) patients still on ventilator at reporting.<br>Overall mortality was 25.8% (56/217), 52 died in ICU, 4 died after transfer out and 40.1% (87/217) patients survived to discharge. | An observational cohort study of all patients with COVID-19 admitted to six COVID-designated intensive care units (ICUs) at three Emory Healthcare acute-care hospitals in Atlanta, Georgia |
| Shekhar et al. [101] | New Mexico USA | 19JAN2020 to 24APR2020 | 50 patients. Median age 55 (range 20 – 85). 27 (54 %) female. Obesity (51 %), diabetes (36 %) hypertension (34 %). 17 direct to ICU, 17 transferred to ICU during stay. 28 of 34 (82 %) required IMV, 5 of 34 HFNC. | 2 | 7 ( in ICU) n= 34<br>20 (ICU, continuing care, n = 6 | 43 of 50 (86 %) were resolved. 24 of 43 (58 %) discharged home; 5 of 43 (11.6 %) to rehab; Mortality 13 of 34 (30.2%) ICU patients died. No non-ICU patient died. | A retrospective and prospective cohort study of the first 50 adults admitted to the University of New Mexico Health Science Centre.<br>7 of 50 continuing care. |
| Crotty et al. [103] | USA Texas | 01MAR2020 to 30APR2020 | 289 Patients. Mean age 59 y (SD 14.4). Hypertension 178 (62 %), diabetes 134 (47 %); 35 coronary heart disease; 35 (12 %); 53 (18 %) no past medical history.<br>241 (83 %) no co-infections; 48 (17 %) co-infection; 25 (8.7% bacterial respiratory co-infection.<br>81 of 289 underwent mech vent for 9, 5 to 14 days. 18 of 25 (72 %) BRC, and 63 of 264 (24 %) no resp co-inf.<br>All hospitalised. 105 of 289 (36.3 %) admitted to ICU. 21 of 25 with bacterial respiratory co-infection admitted to ICU, while 84 of 264 of no resp co-inf (32 %) admitted to ICU. |  | 7.5, 4 – 16.5, all; n = 105<br>8.5, 5 – 12, BRC; n = 21<br>7, 3 – 17, non BRC; n = 84 | 33 of 289 (11.4 %) died. 9 of 25 (45 %) BRC died 24 of 264 (10%) non BRC died.<br>25 of 289 (8 %) continuing care. | A multicentre observational cohort study of SARS-CoV2 positive adult patients age admitted to any of the 4 acute care hospitals within Methodist Health System in Dallas, Texas, USA |
| Kim et al. [104] | USA | 01MAR2020 to 02MAY2020 | 2,491 patients. Median age 62, 50 – 75. 278 of 2,489 (92 %) of patients had ≥1 underlying condition; 1,428 of 2488 (57 %) hypertension, 1,154 of 2332 (50 %) obese. Of those with chronic metabolic disease, 819 of 1024 (80 %) Diabetic.<br>Some 798 of 2,490 (32 %) admitted to ICU; Onset to ICU admission 7, 4 – 10, r 0 – 25 d | 1, 0 – 2, r 0 – 19, n = 798 | 6, 2 – 11, range 1 - 41 n = 771 | 420 of 2490 (17 %) died during hospitalization<br>246 of 462 (53 %) invasive mech vent died. | Cohort study of patients identified through the Coronavirus Disease 2019-Associated Hospitalization Surveillance Network. 154 acute care hospitals in 74 counties in 13 states. |
| **Mexico** |  |  |  |  |  |  |  |
| Valente Acosta et al. [110] | Mexico City | 13MAR2020 to 13APR2020 | 33 patients. Mean age 60.6 ± 12.7 years, 23 (70 %) male. 23 (70 %) overweight or obese. |  | 7.5, 4.25–8.75 Severe n= 20<br>23, 16–24.75 Critical n = 4 | All survived to study cut-off. 20 of 28 severe discharged; 4 of 8 critical discharged | A retrospective case series from a tertiary level private medical centre in Mexico City. 91 % swab positive. All admitted to ICU |

**Figure 5.**
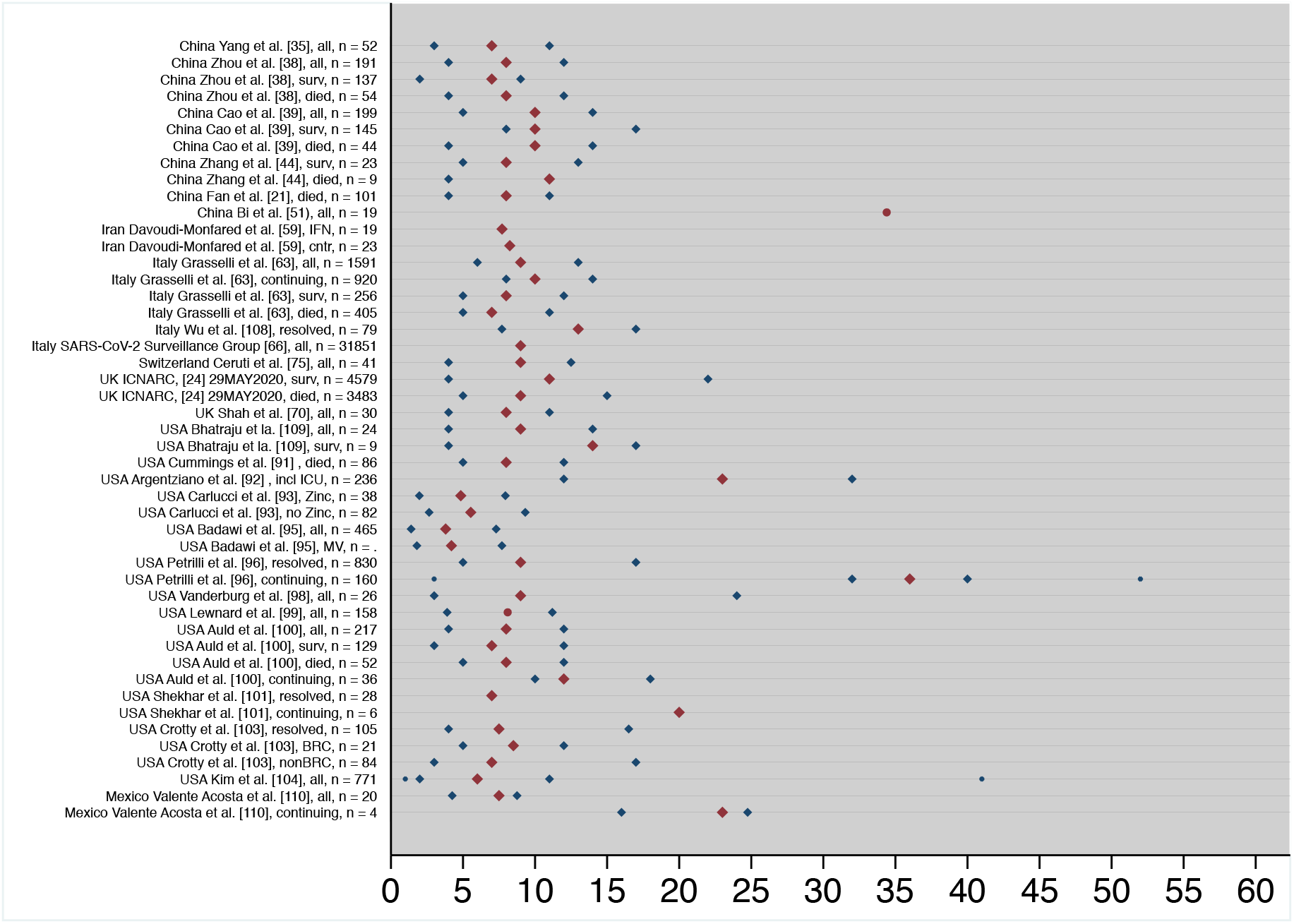
The median (maroon diamond), 25^th^ and 75^th^ percentile (navy diamonds), and range (navy x) for 26 relevant articles and reports describing the median (and IQR) length of stay or mean (maroon circle) in an ICU due to COVID-19 infection. The Italian report (COVID-19 Group, 2020) indicates a median of 9 days (including ICU stay for deceased patients); no estimate of variance was reported. The estimate for Argenziano et al [92] also includes hospital LOS, while Reyes-Gil et al [90] is excluded as it is believed that ICU LOS was calculated with patients who were not admitted to ICU. Studies reporting less than ten patients in ICU [8,57,78,83] were also excluded from Figure 5.

#### Length of hospital stay prior to transfer to ICU

Length of hospital stay prior to transfer to ICU due to COVID-19, by country, is presented in Table 2. Six studies made reference to time in hospital prior to ICU transfer, including two small case series [57,83], thus excluded from consideration. A short LOS prior to ICU admission is outlined in the remainder articles; including 0 days in Fan et al. [21], 1 day in the UK ICNARC report [24] and Kim et al [104], and 2 days in Vanderburg et al [98].

#### Length of hospital post ICU

Length of hospital stay was rarely reported among the studies scrutinized, and thus not described here.

### STUDY MORTALITY RATE

The overall mortality rate for each study is illustrated in Figure 6.

**Figure 6.**
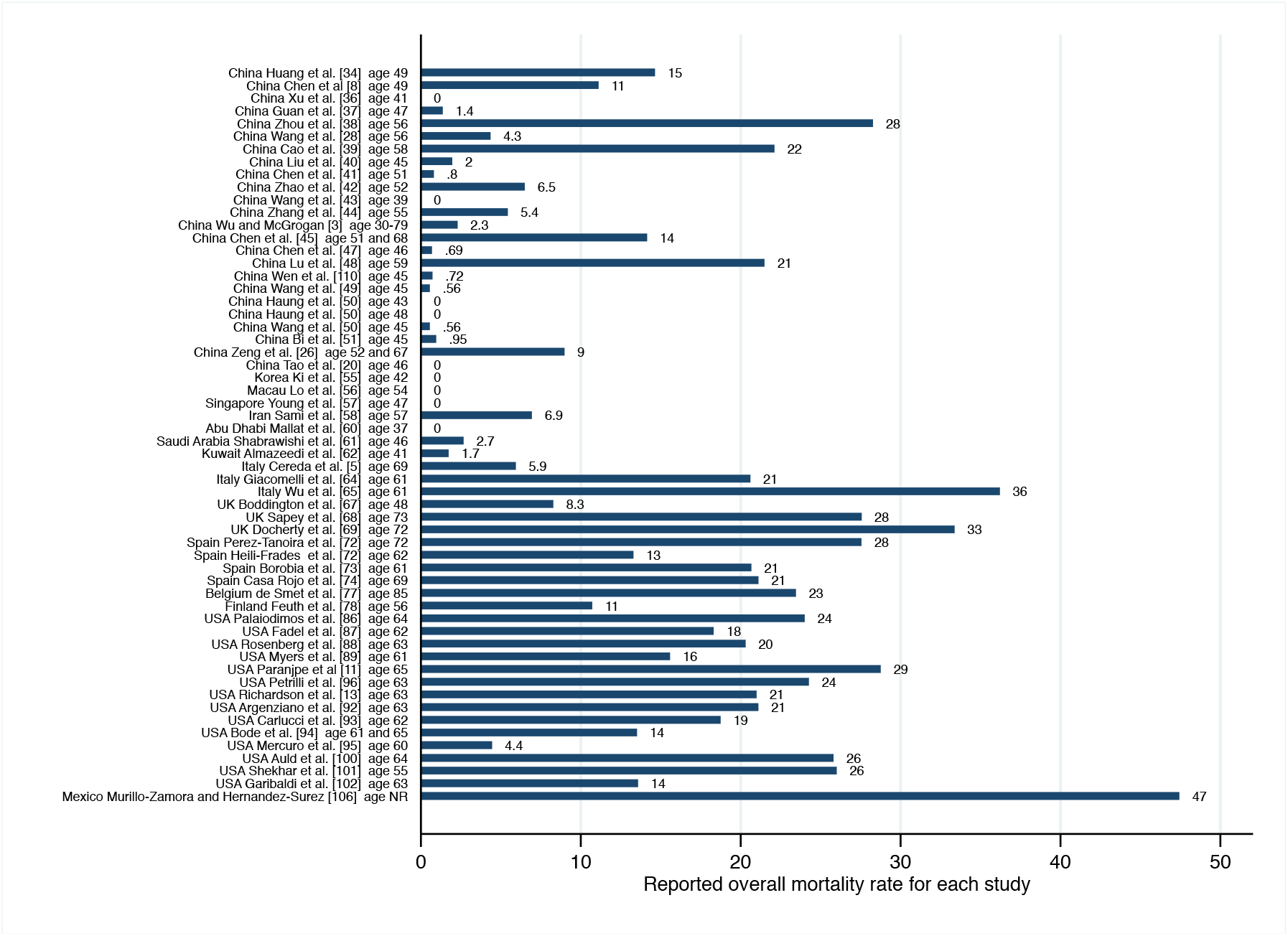
The navy bars indicate the overall mortality rate reported by each study that relates hospitalisation and or hospital length of stay (LOS). The median or mean age reported for study participants is presented. Studies are ordered by country, and by end of enrolment date within country.

## DISCUSSION

### HOSPITALISATION RATE

The relative variation in disease severity is very clear in the published literature from China, with 81 % mild presentation, 14 % moderate presentation with 5 % severe or critical presentations that require high-level on-going care in critical care beds or within the ICU environment [3]. Nevertheless, the experience, thus far, in Europe and the USA would suggest some countries are experiencing a higher degree of severity of COVID-19 being reported. Early indications from Italy suggested hospitalisation rates of 30 % and admission to ICU between 16 to 18 % [5,6]. While the eighth update report from the ECDC [4] restates this but suggests, based on data from 16 EU countries, that respiratory and or ICU support is needed in 2.4 % of cases; the ninth ECDC report [7] slightly revises this country specific median 2 % (IQR 0 to 4 %). Observational studies conducted in the USA also report high rates of hospitalisation [85], high proportion of patients admitted to ICU care and very high mortality rates in hospitalised patients, even among studies with large numbers [11,89,91,91,99,104]. The cause of the differences in disease presentation between countries is not clear. Perhaps differences in case definition, the relative age of the population, the level of pre-existing conditions within the population, and other health factors may be contributing to these differences. Thus far, the experience in Italy suggests that mitigation measures must be carefully managed to ensure ICU capacity is not reached or breached. However, another major influencer on hospitalisation rate will be the definition of a suspect case and the extent of COVID-19 testing within the community; the detection of a greater number of milder cases, diagnosed within the community will ultimately influence the hospitalisation rate. The influence of community testing is becoming evident in Ireland; as April progressed, it was clear in each subsequent HPSC report [79,80,81,82] that the rate of hospitalisation and proportion of patients admitted to ICU was reducing. This is evidence perhaps that a broader case suspect definition and a wider testing approach will impact the overall rate of hospitalisation and admission to ICU.

### LENGTH OF STAY (LOS)

#### Length of stay in hospital (Non-ICU)

The overall length of hospital stay varied between 3 and 52 days among studies. Two of these studies were case series based on the observations of 10 [56] or 12 [83] patients, a further article [22] was based on data extracted from publicly available datasets. The studies are very diverse in their reporting and presentation of hospital length data. The largest study is from Italy [66] reporting the median hospital stay of 5 days for cases that did not require ICU level of care. However, no range or confidence interval is described by this report. Similar to the shorter duration of hospital stay in Italy, a more recent study conducted in the USA [13], which examined hospital records of 5,700 sequential admissions to New York hospitals, reported a short length of stay, with a median 4.1 days (IQR 2.3 to 6.8). The hospital length of stay varies dramatically among different studies. Perhaps differences in hospital capacity, overcrowding, government policy, funding source and other factors might influence length of stay. Of interest Richardson et al. [13] indicates 45 patients were re-admitted to hospital for further care, indicating perhaps the pressure on bed availability. Argenziano et al. [92] outlines the critical situation faced in some Emergency Departments in New York and describes the death of 14.7 % of ED only patients; patients who died in the emergency room prior to being admitted to hospital. Given the large variation in length of stay and the difference between continents; the studies relating to hospital LOS outside an ICU setting were not appropriate to use to calculate a central tendency.

#### Length of stay in hospital (ICU)

With the exception of Bi et al. [51], the majority of Chinese studies reported median ICU stays of 7 to 10 days [21,35,38,39,44]. The Bi et al. [51] study reported exceptionally long hospital and ICU stays, with a median 34 day duration of stay in ICU, for 19 critically ill patients. Similar median lengths of stay have been reported in Italy. The Italian report [66] dated 28^th^ May 2020 describes median LOS of 9 days if admitted to an ICU setting (n = 31,851); however, actual length of time in ICU and proportion or number of patients cared for in ICU were not specified. It is important to note too that this report while including a large number of cases, only includes those who have died, and hence will not reflect the possible longer stay of survivors. Another Italian study conducted by Grasselli et al. [63] reports the median length of ICU stay was 9 days (IQR 6 to 13 days) for 1,581 patients. A Swiss study, too, outlined the median length of stay was 9 days for 41 patients followed to discharge [75]. It is noticeable that length of ICU stay is more variable in US studies. In particular studies that breakdown length of stay into those completed stay compared to patients continuing care. It is evident that care must be taken with interpretation, continuing care groups are reported to have longer stays; median LOS of 11.4 [96]; 14 [109]; and 23 days [92] are outlined. Albeit, the Argenziano et al. study [92] similar to the Italian report [66] reports hospital LOS rather than specific ICU LOS. A smaller case series conducted by Valente-Acosta [110] in Mexico highlights this continuing care group; this study describes a case series of 33 patients admitted to ICU in Mexico city, the median LOS of 23 days is indicative that LOS in ICU can be significant; although their continuing care group is only n = 4. The authors of the UK ICNARC [24] report highlight the continued bias towards shorter stay; with both survivor and non-survivor groups reported to have a median LOS of 11 and 9 days, respectively [24]. Indeed, earlier reports from INCARC [23} reported shorter LOS of ICU patients, as fewer persons had progressed through to completion of treatment. It is evident that study cut-off, and short follow ups, have influenced reported ICU LOS; it is clear that the influence of continuing care groups will lengthen length of ICU stay somewhat in time.

#### Length of hospital stay prior to transfer to ICU

The Median length of hospital stay prior to transfer to ICU was short varying from 0 to 2 days in the relevant articles (21,24,98,104] indicating the short time at which patients not admitted directly to ICU are identified for stepped up care.

### STUDY MORTALITY RATE

It is evident from the data reported in Figure 6 that overall mortality rate varies among studies. In the studies from Asia, the mortality rates were generally low, although rates higher than 10% were noted in 6 studies. In contrast, the mortality rates reported in European and North American studies were substantially higher, in almost all cases exceeding 10% and often much higher. Apparent too, that median (or mean) age differs among these studies; the median age is lower in Asia, only two (of 31) studies report groups with a median age in their sixties, while Wu and Grogan [3] report an overall mortality rate of 2.3 % with 87 % of participants aged between 30 and 79. In contrast, the European and North American studies document an older median age, only one UK study conducted by Boddington et al. [67] reports a median age in the forties (48) for the first few hundred UK COVID-19 cases; indeed 23 of 26 studies conducted in Europe and North America report median age above 60 in Europe of North America. Criteria for hospitalisation will have varied between countries and continents, and are thus likely to influence the relative severity among hospitalised patients. Important too, to note, that some studies such as Bi et al. [51] have clearly indicated that hospitalisation was used as a tool for isolation or quarantine, which is a practice not generally conducted in Europe or North America. An alteration in case definition may also be a factor, and should be further explored. Atypical case presentations for pneumonia are described in the elderly [112,113], and a US review [113] outlines tachypnoea, delirium, unexplained tachycardia or lowered blood pressure as presenting symptoms in the elderly. Early studies from China refer to COVID-19 pneumonia, while more recent studies refer to COVID-19 illness. Differences in case presentation due to age might be a contributor to the relative lower numbers of elderly patients not being included in the studies within China. The influence of pre-existing conditions and BMI within the geographical area also cannot be overlooked.

## CONCLUSIONS

Differences in the severity of case presentations and outcomes have been described between continents, and countries should base its expectation on those cases observed in countries of similar demographics. Advice from the ECDC suggests hospitalisation rates of up to 42 % can be expected; and admission to critical care and ICU levels in up to 3 % of cases, to allow for spare capacity, as mitigation measures are relaxed in our jurisdiction. There is no doubt that the case definition in use and the availability of community testing will determine the hospitalisation rate within a country. In countries where community testing has been significant, we can expect the rate of hospitalisation and the proportion requiring ICU level care to be lower. Data in Ireland certainly supports this assertion.

The length of stay in hospital outside of the ICU setting, due to COVID-19 is highly variable in the literature, with median estimates varying from 3 to 52 days. Studies diverged in their inclusion criterion. It is likely that the aim of hospitalisation varied among countries. Discharge criteria diverged in different jurisdictions, whether it related to a patient required to test negative prior to discharge or where there was pressure to free up overcrowded hospitals. Indeed, it is reported that some countries used hospitalisation to ensure quarantine.

As the pandemic spreads rapidly throughout the world, more and more evidence regarding the length of stay in ICU following COVID-19 has been reported. Many of these reports indicate a median ICU length of stay between 7 to 11 days; however a number of these studies report length of stay based only on resolved cases, and as such duration of stay is likely skewed towards a short stay due to bias towards those who have completed their ICU stay and are now discharged or have died. Thus the upper estimates of the range should be considered for use in resource models. It is apparent too that the data generated by each individual country during the initial months of the outbreak will be critical to estimate the parameters used for any national resource model.

## Data Availability

Data can be made available on request.

